# Multiparametric MRI Along with Machine Learning Informs on Molecular Underpinnings, Prognosis, and Treatment Response in Pediatric Low-Grade Glioma

**DOI:** 10.1101/2024.04.18.24306046

**Authors:** Anahita Fathi Kazerooni, Adam Kraya, Komal S. Rathi, Meen Chul Kim, Arastoo Vossough, Nastaran Khalili, Ariana Familiar, Deep Gandhi, Neda Khalili, Varun Kesherwani, Debanjan Haldar, Hannah Anderson, Run Jin, Aria Mahtabfar, Sina Bagheri, Yiran Guo, Qi Li, Xiaoyan Huang, Yuankun Zhu, Alex Sickler, Matthew R. Lueder, Saksham Phul, Mateusz Koptyra, Phillip B. Storm, Jeffrey B. Ware, Yuanquan Song, Christos Davatzikos, Jessica Foster, Sabine Mueller, Michael J. Fisher, Adam C. Resnick, Ali Nabavizadeh

## Abstract

In this study, we present a comprehensive radiogenomic analysis of pediatric low-grade gliomas (pLGGs), combining treatment-naïve multiparametric MRI and RNA sequencing. We identified three immunological clusters using XCell enrichment scores, highlighting an ‘immune-hot’ group correlating with poorer prognosis, suggesting potential benefits from immunotherapies. A radiomic signature predicting immunological profiles showed balanced accuracies of 81.5% and 84.4% across discovery and replication cohorts, respectively. Our clinicoradiomic model predicted progression-free survival with concordance indices of 0.71 and 0.77 in these cohorts, and the clinicoradiomic scores correlated with treatment response (p = 0.001). We also explored germline variants and transcriptomic pathways related to clinicoradiomic risk, identifying those involved in tumor growth and immune responses. This is the first radiogenomic analysis in pLGGs that enhances prognostication by prediction of immunological profiles, assessment of patients’ risk of progression, prediction of treatment response to standard-of-care therapies, and early stratification of patients to identify potential candidates for novel therapies targeting specific pathways.

## 1 Introduction

Pediatric low-grade glioma (pLGG), classified as grade I or II tumors by the World Health Organization (WHO), constitutes the most common childhood brain tumors ^1,2^. They represent roughly 30% of all pediatric central nervous system tumors and include a diverse spectrum of histologies, from glial to mixed glioneuronal entities ^1,2^. The prognosis and treatment responses among patients with pLGG vary widely. Gross total resection offers the most favorable response in pLGG patients with 10-year progression-free survival (PFS) exceeding 85% ^3^. Complete resection cannot be achieved for all tumors, especially for highly infiltrative or deep-seated tumors. Incompletely resected tumors often require additional therapy ranging from conventional chemotherapy to targeted inhibitors. Notably, for a significant portion of patients, the tumor progresses, resulting in a 10-year PFS rate under 50%, leading to concerns about long-term complications and an elevated mortality risk ^3^.

The potential of molecularly targeted therapeutics to transform pLGG treatment is increasingly recognized, as evidenced by the FDA approval and ongoing clinical trials of RAF and MEK inhibitors ^4,5^. Nonetheless, a thorough understanding of the biological and molecular foundations of pLGG tumors, beyond merely identifying specific markers such as *BRAF* alterations, is crucial to the success of targeted treatments. This knowledge is also essential in preventing paradoxical tumor growth as seen with the sorafenib kinase inhibitor drug in *BRAF*-altered pLGGs ^4^. Accordingly, with increasingly encouraging reports on the efficacy of immunotherapies in a number of solid pediatric brain tumors ^6^, it is vital to understand the immunological characteristics of brain tumors. The tumor immune microenvironment (TIME) – composed of a diverse set of immune and stromal cells exhibiting complex interactions with each other and with tumor cells – influences the suppression or promotion of tumor growth. It plays a significant role in shaping the prognosis and affecting the response to treatment ^7^.

Radiomics has demonstrated potential in offering non-invasive, *in vivo* biomarkers of tumor molecular composition, which complement well-established clinical variables in supporting risk stratification by quantifying the underlying tumor heterogeneity and revealing the molecular underpinnings of the tumor via radiographic phenotypes (i.e., radiophenotypes) ^8^. We propose that in pLGG, the use of radiomic analysis on treatment-naïve multiparametric MRI (mpMRI) sequences coupled with machine learning (ML) can reveal associations between radiophenotypes and the tumor’s molecular composition, encompassing the TIME and prospective treatment targets. This approach extends the scope of analysis beyond the capabilities of molecular subtyping, which has traditionally been the focus of radiogenomic research in pLGGs ^9–12^. By integrating imaging-derived phenotypes of pLGG with genotypic traits discerned from transcriptional analysis, our goal is to offer an in-depth characterization of pLGGs, aiding the implementation of precision therapies.

This study provides a comprehensive radiogenomic analysis through creating a model for characterization of pLGG tumor immune microenvironment (TIME) subgroups, developing a radioimmunomic signature for predicting these immunological subgroups, and generating a clinicoradiomic model for predicting pLGG progression-free survival, assessing progression risk and treatment response. Additionally, an interpretability analysis was conducted to identify pathways associated with the predicted risk.

## 2 Results

### 2.1 Study Design and Cohort Description

The study design focuses on three main elements (Figure 1): (1) predicting pLGG immunological profiles, (2) predicting the immunological subgroups using a radioimmunomic signature, (3) developing a clinicoradiomic model to predict progression-free survival, along with an interpretability analysis of transcriptome pathways that are related to the risk of tumor progression. We analyzed retrospective data from 545 patients (See Table 1 for detailed description) in the Children’s Brain Tumor Network (CBTN ^13^), encompassing 494 with multi-omic (RNA-seq and WGS) data and 201 with standard multiparametric MRI (mpMRI) scans. A subset of 150 had both WGS and mpMRI data, with 91 also including diffusion-weighted imaging (and apparent diffusion coefficient (ADC) maps). The imaging cohort (201 patients) was split into a ‘discovery’ (160 patients) and an unseen ‘replication’ (41 patients) cohort randomly, ensuring the inclusion of rare tumor locations like the basal ganglia in the discovery group. This division was consistent in all ML models, although sample sizes varied for analyses combining transcriptomic data. Specifically, the radioimmunomic analysis included 120 patients in the discovery group and 30 in the replication set.

**Figure 1.**
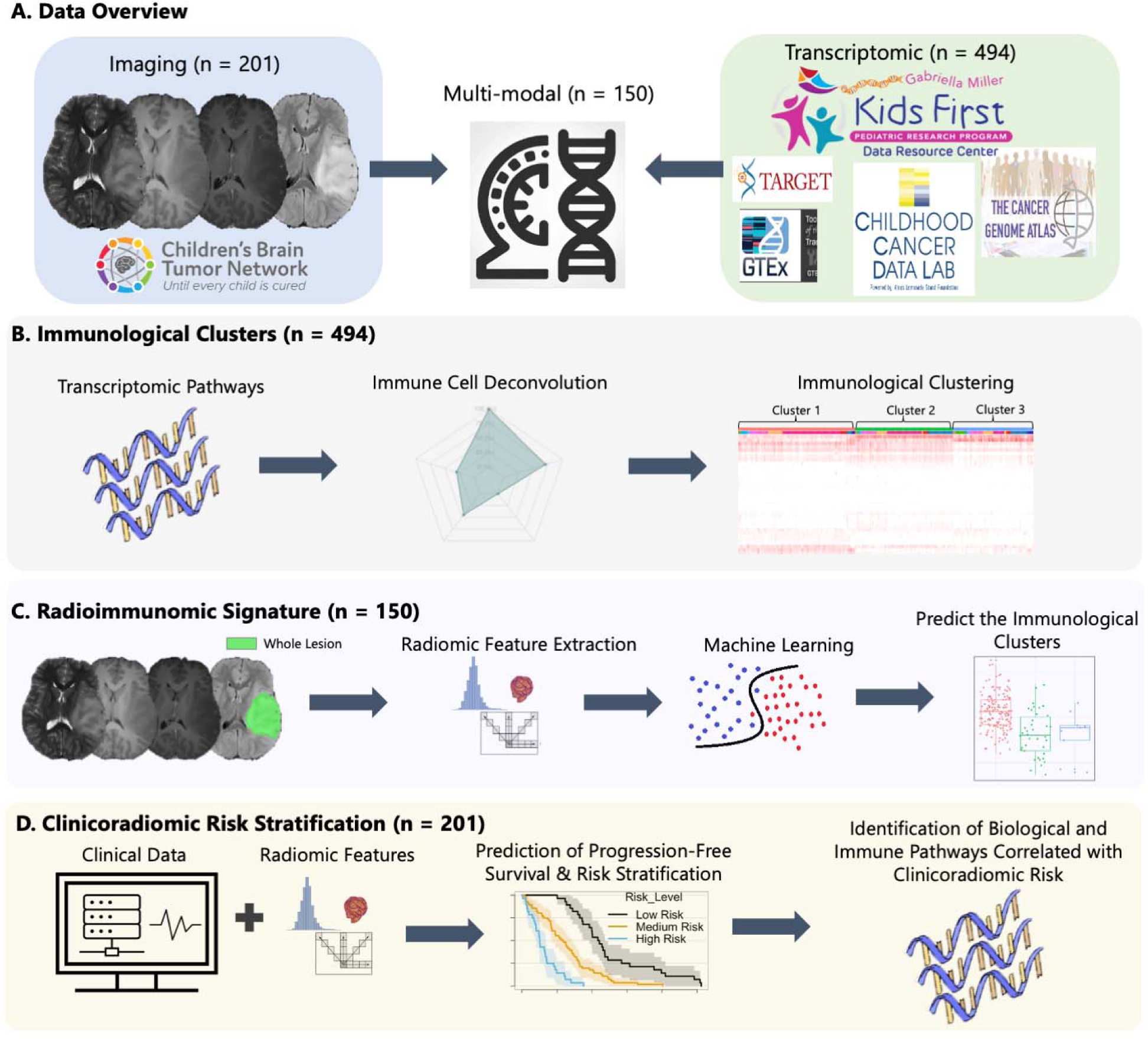
Graphical description of the primary analyses in this study: (A) Overview of the imaging, transcriptomic, and multi-modal (combination of imaging and transcriptomic) cohorts. (B) Identification of the immunological clusters within pediatric low-grade glioma. (C) Generating a radioimmunomic signature to predict immunological clusters. (D) Developing a clinicoradiomic model that incorporates radiomic features and clinical data to predict progression-free survival in pLGG and thereby, patients’ risk of progression, stratify risk, and analyze the biological and immune pathways linked to clinicoradiomic risk.

**Table 1.**
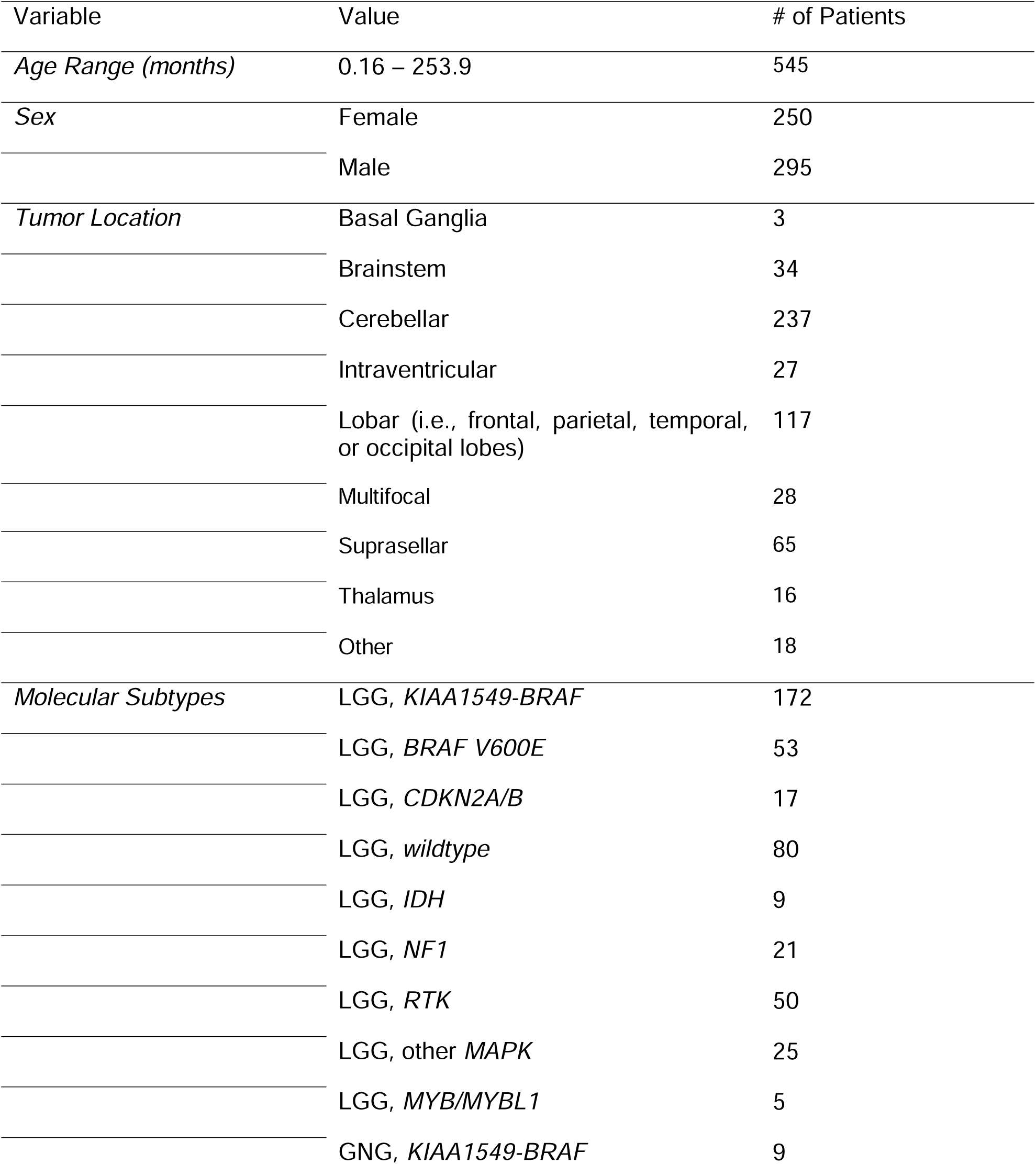
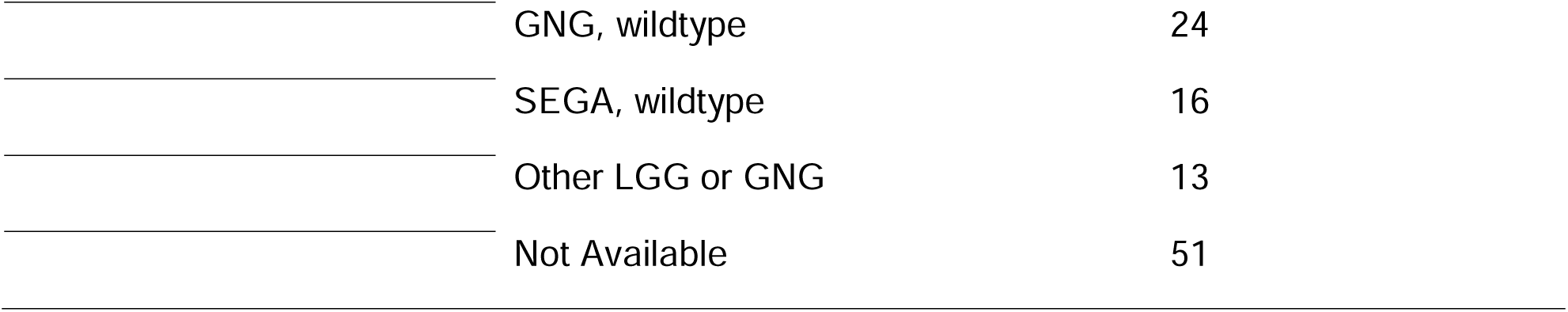
Summary of demographics and clinical characteristics of the pLGG patients included in this study (collected through the CBTN data repository).

### 2.2 Transcriptome Enrichment Analysis Identifies Three Immunological Clusters

Clustering of 494 pLGGs with available RNA-sequencing data in our cohort revealed three distinct immunological groups based on immune cell infiltration-related gene expression, with immune cluster 1 (n = 189) showing intermediate inferred infiltration, immune cluster 2 (n = 164) showing high inferred infiltration, and immune cluster 3 (n = 141) showing the lowest inferred immune cell infiltration (Figure 2A). We observed a significant negative correlation between immune score and estimates of tumor purity derived from copy number (ABSOLUTE) ^14^, expression (ESTIMATE) ^15^, and methylation-based assessments (LUMP) ^16^ (Supplemental Figure 1).

**Figure 2.**
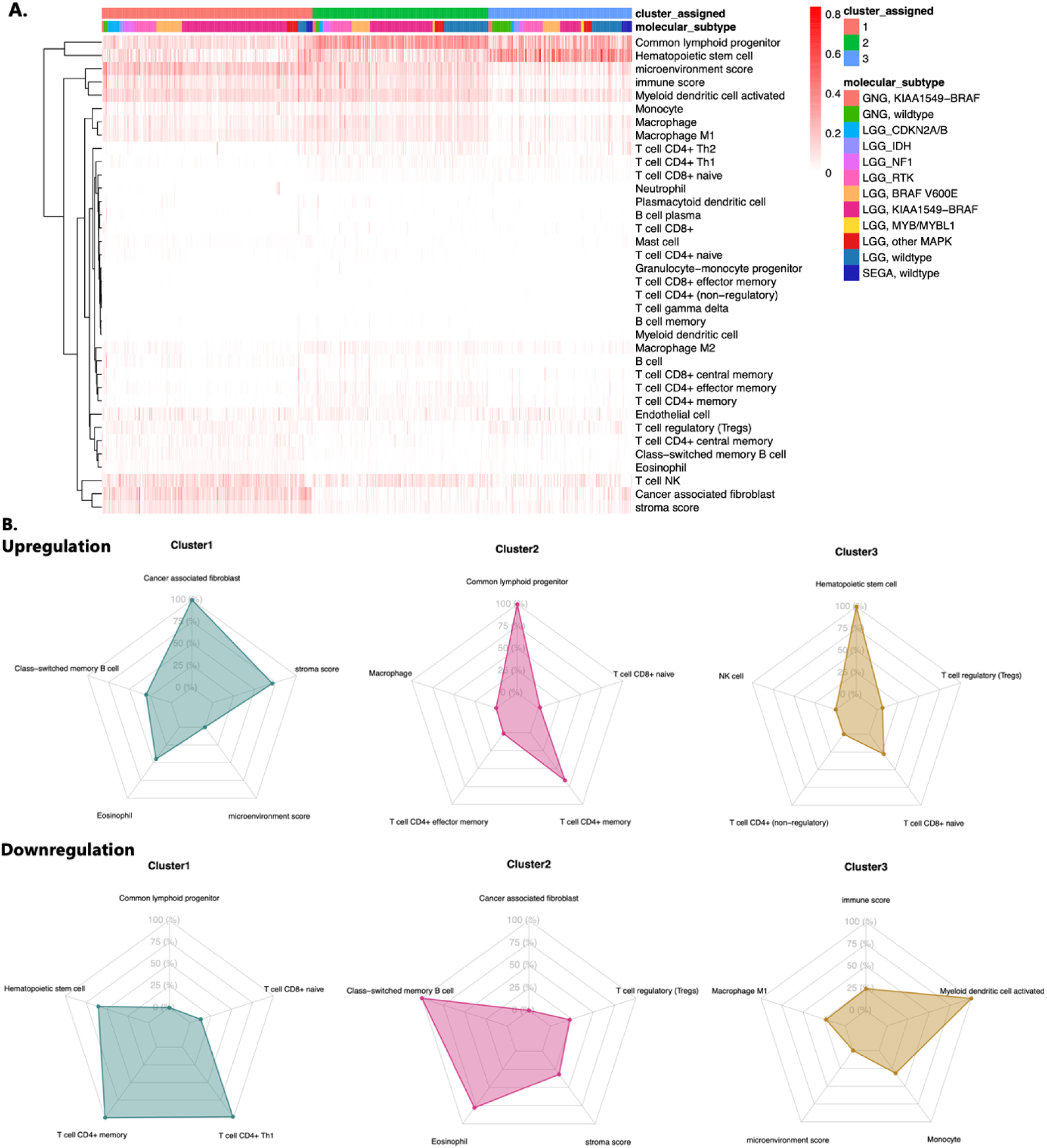
(A) Heatmap indicating the expression levels of immune cells across different patients in the three identified immunological profiles using XCell method (indicated above the heatmap as “cluster assigned”, along with the pLGG molecular subtypes for this cohort of subjects. (B) Radar plots indicating the upregulated and downregulated set of cell types in each of the immunological clusters.

Cluster 1 showed highest enrichment in stromal scores, eosinophils, and cancer-associated fibroblasts (CAFs) and lower levels of hematopoietic stem cells (HSCs), memory CD4+ T-cells, and CD4+ Th1 cells than clusters 2 and 3 (Figure 2B). Cluster 2 illustrated higher levels of common lymphoid progenitors, M2-polarized macrophages, and CD4+ Memory T-cells and the lowest enrichments for class-switched memory B-cells, eosinophils, T-regulatory cells (Tregs), and stromal scores compared to clusters 1 and 3 (Figure 2B). Cluster 3 exhibited higher expression of hematopoietic stem cells (HSCs), Tregs, and naïve CD8+ T-cells and lower levels of activated myeloid dendritic cells, monocytes, and M1-polarized macrophages in comparison with clusters 1 and 2 (Figure 2B).

We also found that expression of the tumor inflammation signature (TIS), a clinically predictive expression-based measure of response to nivolumab and pembrolizumab ^17^, was statistically significant between clusters 1 and 2 (p = 2.6e-6), clusters 1 and 3 (p = 2.4e-8), and clusters 2 and 3 (p = 2.2e-16) (Figure 3 C & D). Cluster 2 revealed the highest TIS across all the immune groups, consistent with the independent measures of immune infiltration-related genes that we observed from XCell ^18^ with this group. Notably, tumor mutational burden (TMB), a clinical biomarker of response to immune checkpoint blockade ^19,20^, was significantly different between Cluster 1 and Cluster 2 (p = 3.9e-5) and between Cluster 1 and Cluster 3 (6.3e-5), but not between Clusters 2 and 3 (p = 0.75) (Figure 3E).

**Figure 3.**
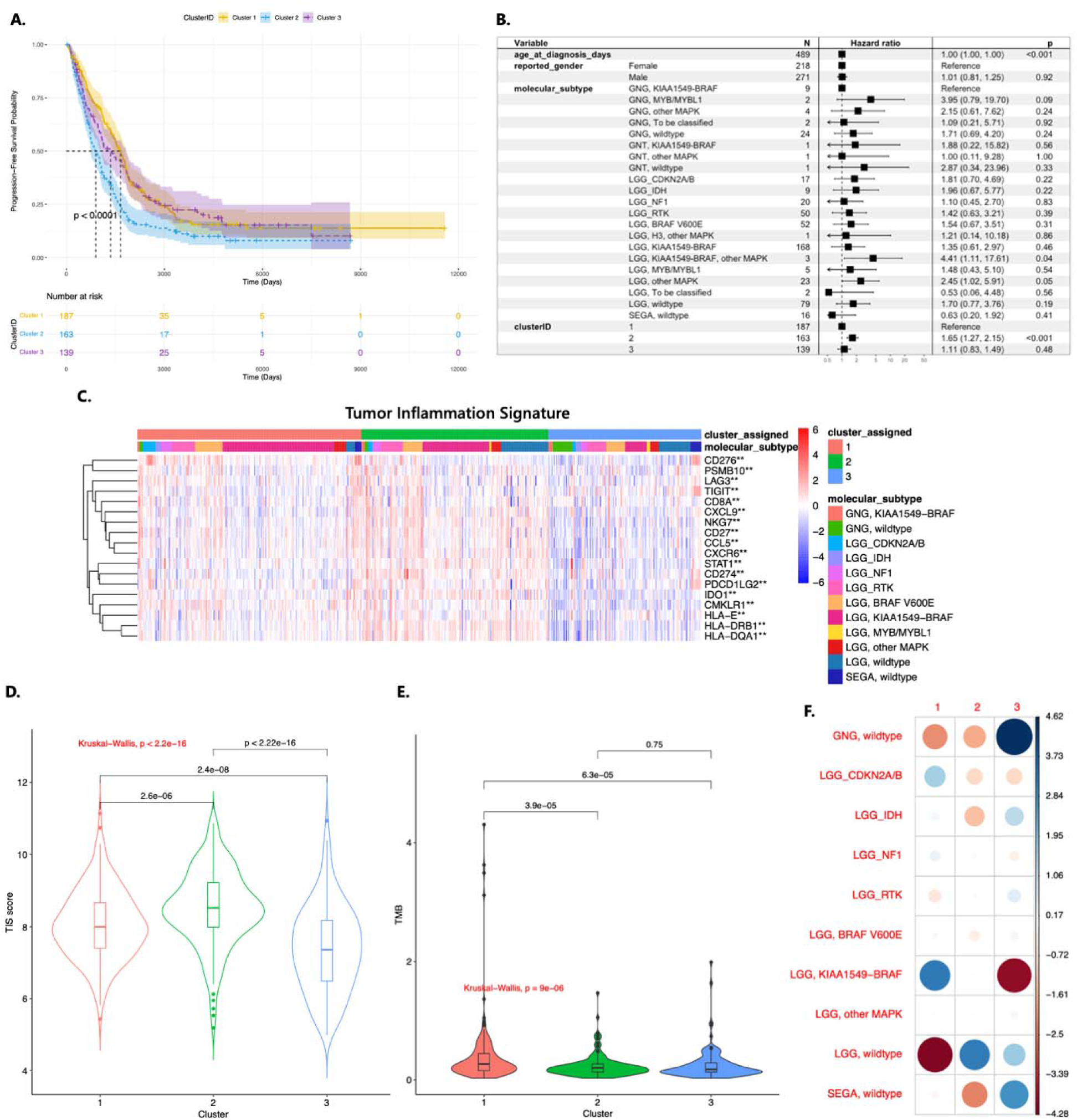
(A) Kaplan-Meier curves representing the probability of progression-free survival across different immunological clusters, suggesting a statistically significant difference (p<0.0001) between the clusters based on log-rank comparison. (B) Forest plot from Cox regression analysis illustrating the effects of immunological cluster assignment, molecular subtypes, age, and sex on PFS. (C) The heatmap of tumor inflammation signature (TIS) as a composite of 18 genes. The association with the molecular subtypes and Xcell clusters is indicated at the top of the heatmap plot. (D-E) Violin plots representing the distribution of the values for TIS and tumor mutational burden (TMB), respectively, across the three immunological clusters. (F) Matrix of Pearson residuals illustrating the relationship between immune groups and pLGG molecular subtypes. Circle sizes are proportional to the Pearson residual values. Shades of blue and red, with varying degrees of opacity, indicate the degree of overrepresentation or underrepresentation of specific molecular subtypes within each immune cluster.

We evaluated the relationship between immune groups with known pLGG molecular subtypes related to WHO brain tumor entities using a chi-square test of independence to ascertain a potential association between immunophenotype and molecular driver events (Figure 3F). To delineate specific XCell cluster and molecular subtype combinations that deviated from expectation, we quantitatively determined significant group-level associations through a Poisson generalized linear model. The LGG “wildtype” subtype, which refers to tumors that do not harbor a driver mutation in Histone *H3*, *IDH*, *FGFR1/2* genes, or MAPK pathway components (*KRAS, NRAS, HRAS, BRAF MAP2K1, MAP2K2, ARAF, or RAF1*), was underrepresented in the immune-high cluster 1 (Figure 3F), while tumors harboring a *KIAA1549::BRAF* fusion, partial or complete losses in *CDKN2A/B*, MAPK pathway mutations, or mutations in *IDH*, *NF1*, or receptor tyrosine kinases (RTKs, *MET, KIT, PDGFRA, ALK, ROS1, NTRK1/2/3)* were over-represented in this group (Figure 3F, Supplemental Figures 4 -5, p < 0.001). Wildtype subependymal giant cell astrocytomas (SEGAs) were also under-represented in this cluster (Figure 3F, Supplemental Figures 4 - 5, p < 0.001). In cluster 2, which showed a similar level of immune infiltration-related gene expression overall relative to cluster 1 based on the XCell immune score, we observed the reverse trend among LGG wildtype tumors, which were over-represented (Figure 3F). By contrast, wildtype GNGs (p < 0.001) and tumors harboring *IDH* mutations (p = 0.0496), tumors with *BRAF* alterations, and tumors harboring mutations in RTK genes were under-represented in this immune group (Figure 3F, Supplemental Figures 4 - 5, p < 0.001). In the immune-low cluster 3, we observed strong over-representation of wildtype GNGs and SEGAs and a strong under-representation of *KIAA1549::BRAF* fusion-driven pLGGs from an analysis of the Pearson residuals in Figure 3F. Collectively, these data suggest that tumor-cell intrinsic factors may be important mechanistic determinants of the tumor-immune phenotype in pLGGs, an observation that has also been noted in a number of other tumor types ^21–23^. These data, along with our observations of associations between immune clusters with TIS and TMB, suggest that subtype-specific driver mutations may be more important and specific determinants with respect to the tumor-immune microenvironment and that total mutational count may play a secondary role given our observations between Clusters 2 and 3 ^19,20^.

We then evaluated the association of the three immunological clusters with prognosis using survival endpoints progression-free survival (PFS) and overall survival (OS) (Figure 3 A&B; Supplemental Figure 2). A log-rank comparison across the three clusters revealed that the immune-high cluster 2 had the worst overall prognosis, followed by cluster 1 which illustrated intermediate infiltration-related gene expression, and finally cluster 3, which was predicted to have a low level of immune cell infiltration. Notably, the cluster 2 subgroup had the highest expression of TIS and cluster 3 the lowest, and TIS has been shown to inversely associate with survival in adult low-grade gliomas consistent with our observations ^24^. When modeling the effects of cluster when adjusted for age at diagnosis, reported sex, race, the anatomical region of the tumor, and the molecular subtype of the tumor in a Cox regression model, we found significantly higher progression-related hazard for tumors belonging to cluster 2 (p < 0.001, Figure 3B; Supplemental Table 2). The OS outcomes were not statistically different among the three patient groups by non-parametric Kaplan Meier analysis (Supplemental Figure 2).

We evaluated the relationship between immunological clusters and tumor locations, investigating if specific brain regions are inclined to exhibit distinct immune microenvironments. As the Sankey diagram in Supplemental Figure 6 presents, the immunological clusters are distributed fairly evenly across the cerebellar region, which is the most common tumor location across all three immunological clusters, followed by lobar tumor location (i.e., frontal, parietal, temporal, or occipital lobes). Suprasellar tumors were predominantly associated with immune-high clusters (1 and 2) and were less common in the immune-low cluster 3. This pattern indicates that there is no significant predilection for specific brain regions for immunological profiles.

### 2.3 Radioimmunomic Signature: Differentiating Between Poor and Favorable Prognosis Immune Clusters

We assessed the potential of radiomic analysis in predicting pLGG immunological clusters, focusing on developing a radioimmunomic signature using machine learning classifiers. Since immune clusters 1 and 3 were not prognostically distinct (Figure 3A), we aimed to distinguish cluster 2 from clusters 1 and 3. Our analysis involved 150 patients with conventional MRI sequences and 91 with additional ADC-map data. We trained radioimmunomic models on imaging features and age for prediction, testing them independently on the replication set. The conventional MRI-based radioimmunomic signature for cluster 2 versus clusters 1 and 3 showed an AUC of 0.77 | 0.74 and balanced accuracy of 76.8% | 86.0% in the discovery | replication sets. Adding ADC features to conventional MRI improved performance, with an AUC of 0.83 | 0.79 and balanced accuracy of 81.5% | 84.4% in the discovery | replication sets (Figure 4 A).

**Figure 4.**
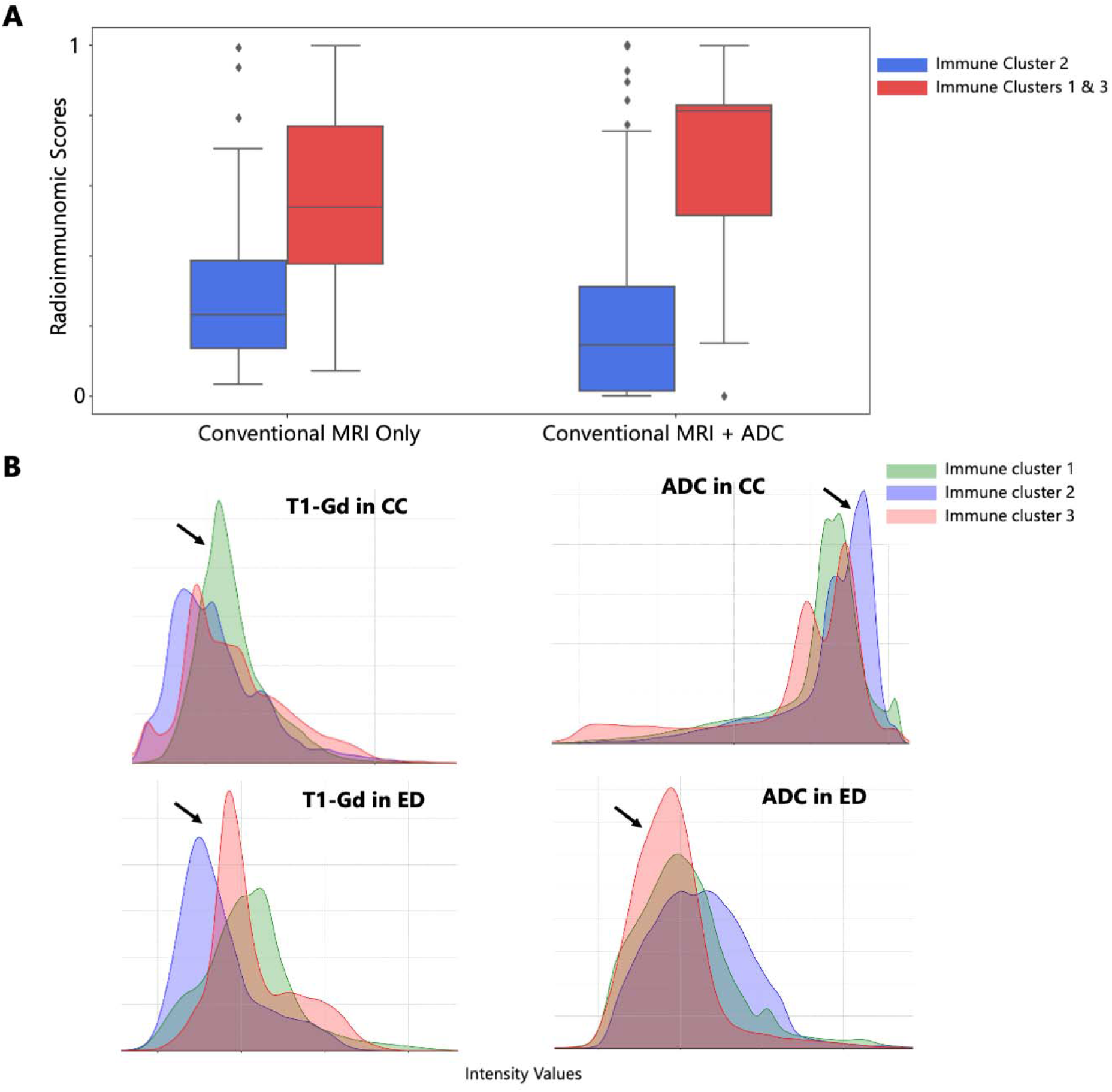
(A) Radioimmunomic signatures generated for classification of Immune Cluster 2 versus Clusters 1 and 3, based only on conventional MRI features, as well as conventional MRI + ADC features (B) Histograms of image intensities within various tumorous subregions for the tumors in immunological clusters 1, 2, 3.

Our analysis further demonstrated different imaging characteristics within tumor subregions across the three immunological clusters (Figure 4 B). Immune cluster 2 exhibits more pixels with low enhancement in the ED tumor subregions, while cluster 3, characterized by lower immune cell expression and higher tumor purity (section 2.1.1 and Supplemental Figure 1), showed more pixels in the lower ADC range in the ED region. This possibly indicates restricted diffusion and a higher tumor cell density rather than immune cell infiltration in peritumoral edema. A negative correlation between ADC values and tumor purity within the peritumoral edema has been reported in adult high-grade glioma ^25^.

### 2.4 Predictive Clinicoradiomic Model: Assessing Progression-Free Survival and Treatment Resistance Association

We hypothesized that a combination of clinical variables and radiomic features from pre-surgical/pre-treatment MRI sequences could predict tumor progression risk. In our discovery cohort (n = 160), we developed a Cox proportional hazards model combining these variables. Ridge regularization was applied to radiomic variables, while clinical variables remained unpenalized (refer to Methods section 5.4). This model, tested on the replication set, aimed to predict patient risk scores (Figure 5A). In the discovery and replication sets, the clinicoradiomic model achieved Harrell’s concordance indexes of 0.71 [95% CI: 0.63, 0.79] and 0.77, Uno’s concordance indexes of 0.72 [95% CI: 0.64, 0.80] and 0.80, and integrated Brier scores (IBS) of 0.24 [95% CI: 0.19, 0.29] and 0.16, respectively (Table 2). Supplemental Figure 7 shows hazard ratios for all variables in ascending order. Key clinical predictors include tumor resection extent, chemotherapy, age at diagnosis, and tumor location. Regularization selected 22 radiomic features, with 8 statistically significant. Patients were categorized into low, medium, and high-risk groups, with risk cutoffs applied independently in the replication set. ANOVA tests yielded p-values of 3.12e-58 and 1.29e-54 across risk groups in the discovery and replication sets.

**Figure 5.**
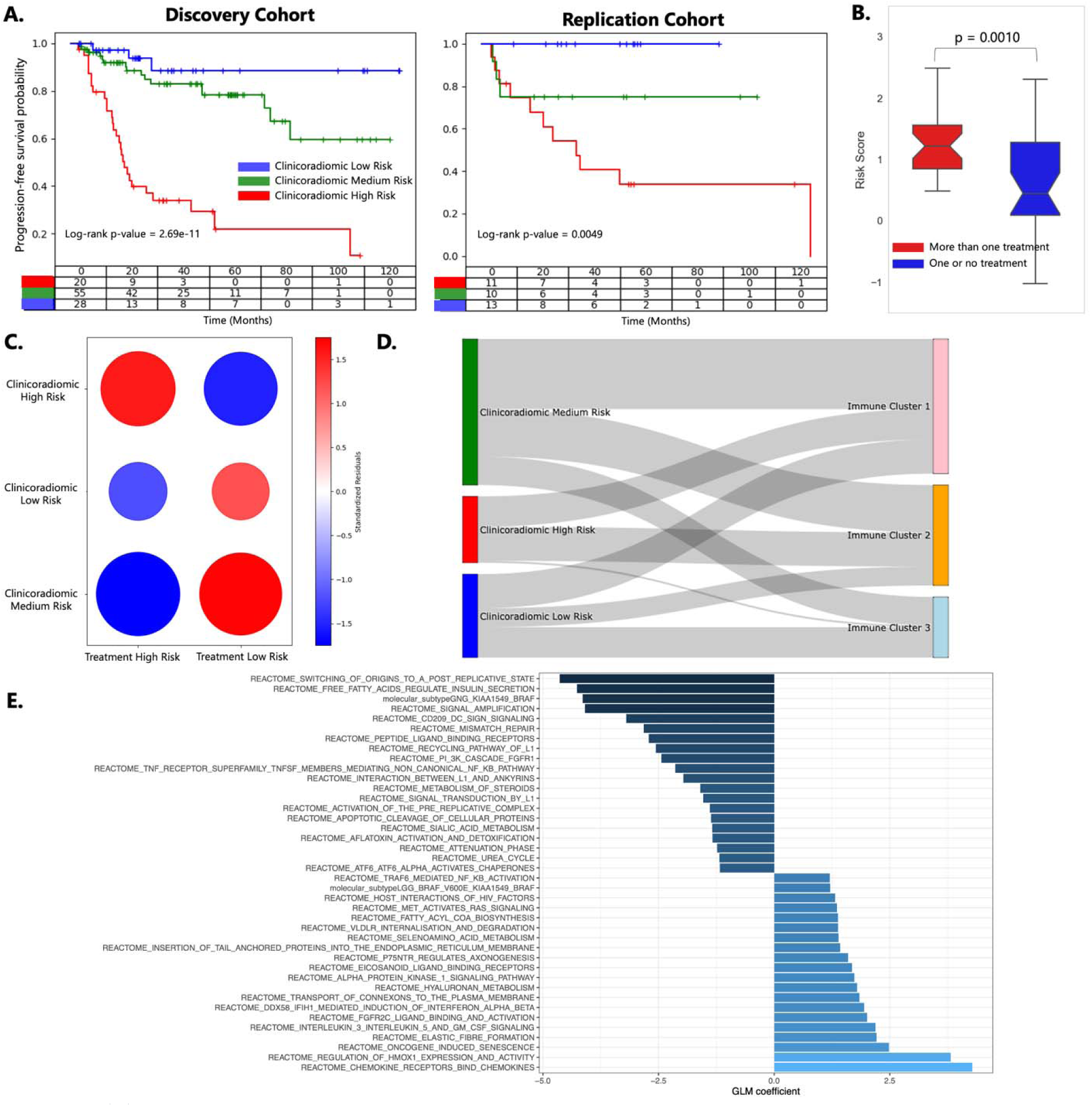
(A) Kaplan-Meier Curves for the Progression-Free Survival Probability of Clinicoradiomic Risk Scores in the Discovery and Replication cohorts. (B) Box-and-Whisker plots indicating the distribution of risk scores in the patients that received one or no “systemic” treatments (low-risk) (see section 2.3.1 in the manuscript for the definition of the two treatment risk categories), compared to those who received more than one treatment (high-risk) over the course of five years. (C) Association plot of standardized residuals for the relationship between the clinicoradiomic and the treatment risk categories (the numbers within each circle in the association plot represent the standardized residuals from the chi-square test of independence). Plots B-C only present the data for the subjects that had progression over a course of five years (n = 61). (D) Sankey plot illustrating the association between the Clinicoradiomic risk groups and the immunological clusters. (E) Bar plot of GLM coefficient derived from an elastic net regression model illustrating pathways associated with the predicted clinicoradiomic risk.

**Table 2.** Summary of the performances of clinical-only and clinicoradiomic models on all subjects, and a subgroup analysis on patients who did not undergo gross/near-total resection. The performance metrics include Harrell’s concordance index (HAR), Uno’s concordance index (UNO), and the Integrated Brier Score (IBS)

|  | **HAR** | **UNO** | **IBS** |
| --- | --- | --- | --- |
| **Model 1: Clinical-Only** |  |  |  |
| Discovery | 0.74 [95% CI: 0.67, 0.80] | 0.75 [95% CI: 0.67, 0.82] | 0.20 [95% CI: 0.16, 0.22] |
| Replication | 0.76 | 0.79 | 0.16 |
| **Model 2: Clinicoradiomic** |  |  |  |
| Discovery | 0.71 [95% CI: 0.63, 0.79] | 0.72 [95% CI: 0.64, 0.80] | 0.24 [95% CI: 0.19, 0.29] |
| Replication | 0.77 | 0.80 | 0.16 |
| **Subgroup Analysis (Subjects who did not undergo gross/near-total resection)** |  |  |  |
| Model 1: Clinical-only | 0.55 | 0.57 | 0.24 |
| Model 2: Clinicoradiomic | 0.70 | 0.70 | 0.18 |

Our analysis revealed that a model based solely on clinical variables did not yield markedly different results from those of the clinicoradiomic model, as detailed in Table 2 and Supplemental Section 6.3. Nevertheless, given the notable predictive importance of total resection in pLGGs, we compared the efficacy of the trained clinicoradiomic and clinical-only models specifically in patients who did not undergo total/near-total resection. In this subset, the clinicoradiomic model demonstrated significantly enhanced performance relative to the clinical-only model, as evidenced by higher Harrell’s concordance indexes (0.70 vs 0.55), Uno’s concordance indexes (0.70 vs 0.57), and a lower IBS of 0.18 compared to 0.24 for the clinicoradiomic and clinical-only models, respectively (Table 2). This finding underscores the clinicoradiomic model’s potential value in stratifying patients who are not adequately classified using only clinical variables.

We investigated the relationship between our predicted risk scores and treatment resistance, focusing on patients who showed progression (n = 61) within five years post-diagnosis. These patients were divided into two groups based on their treatment: those receiving minimal systemic therapy (chemotherapy or, rarely, radiotherapy post biopsy/surgery; n = 31), forming a treatment low-risk group, and those undergoing multiple treatments (n = 30), forming a treatment high-risk group. A Student’s t-test revealed a significant difference in mean predicted risk scores between these groups (p = 0.0010), with higher risk scores linked to patients less responsive to initial treatments, as shown in Figure 5B.

Our analysis of the clinicoradiomic risk groups in relation to treatment risk categories revealed a significant association, indicated by a Chi-squared test statistic of 13.69 and a p-value of 0.0011. As Figure 5C shows, there is an over-representation of clinicoradiomic high-risk patients in the treatment high-risk group, and an under-representation in the treatment low-risk group. Conversely, clinicoradiomic low-risk patients are more prevalent in the treatment low-risk group and less so in the treatment low-risk group. Most medium-risk clinicoradiomic patients fall into the treatment low-risk category. These findings imply that patients with higher clinicoradiomic risk scores may require multiple lines of treatment.

We explored correlations between clinicoradiomic risk categories and immunological clusters. Figure 5D illustrates this relationship, showing a rare occurrence of immune cluster 3 in the clinicoradiomic high-risk group, and a high occurrence of the prognostically worse immune cluster 2 in the clinicoradiomic high or medium-risk groups. Most of the tumors in immune cluster 1 were grouped in the medium and low-risk groups.

Our findings show that immune cluster 3, linked to a favorable prognosis, rarely appears in the predicted high-risk category, validating our clinicoradiomic model’s ability to predict immunological factors related to progression risk. Predominantly, the clinicoradiomic high-risk group includes patients from immunological clusters 1 and 2, characterized by an inflamed yet possibly tumor-promoting immune environment.

Finally, we examined how molecular subtypes enhance the clinicoradiomic model’s accuracy in predicting tumor progression risk. This analysis yielded Harrell’s concordance indexes of 0.74 [95% CI: 0.65, 0.83] and 0.69, Uno’s concordance indexes of 0.74 [95% CI: 0.65, 0.84] and 0.74, and integrated Brier scores (IBS) of 0.24 [95% CI: 0.14, 0.27] and 0.18, for the discovery and the replication sets, respectively, as detailed in Supplemental Material, section 6.3.3.

### 2.5 Biological Interpretability of Clinicoradiomic Risk Scores: Associations with Germline Variants and Transcriptomic Pathways

We then aimed to independently investigate whether genomic and transcriptomic characteristics may be associated with radiomically-defined risk of progression. We interrogated deleterious and likely deleterious germline variants present in the pLGG population through the criteria outline in Materials and Methods (Section 4.1.2.3), annotating for presence or absence of a germline variant among cancer genes and those involved in structural birth defects ^26–29^ (Supplementary Table 7). From a chi-square test of independence testing for the association of gene-specific germline variant counts across clinicoradiomic risk groups, we identified the synaptic nuclear envelope protein 1 (*SYNE1*) gene as significantly over-represented in the medium risk clinicoradiomic group (adjusted p = 0.015). Notably, *SYNE1* germline mutations have been implicated in cerebellar ataxia and somatic mutations of the gene are known to contribute to the development and progression of multiple cancers, among them adult glioblastoma ^30–33^.

We performed single-sample GSEA (ssGSEA) to derive per-sample enrichment scores for 1692 Reactome pathways spanning across all known cellular biological functions. Consistent with prior studies ^23,34^, we found that the *KIAA1549::BRAF* fusion subtype was linked to lower risk of progression. Among notable pathways negatively associated with risk and cancer initiation/progression, we identified CD209+ dendritic cells, with an important role in initiating ^35^ and non-canonical NFKB signaling ^36–38^. Furthermore, we found that up-regulated signaling of the eicosanoid family of lipid mediators, produced by poly-unsaturated fatty acids, were associated with higher risk, consistent with their known pro-angiogenic, proliferative, and inflammatory effects in glioma ^39–41^. We identified several inflammatory pathways with known biological significance in glioma that were associated with greater risk. Interleukin-3, suppressing anti-tumor immune responses via activation of PDL1 pathway ^42^, and Interleukin-5 signaling that promotes cancer metastasis by remodeling the TIME ^43^, were associated with greater risk of progression. Type I interferon signaling, previously found to promote a mesenchymal phenotype and poor prognosis in glioma ^44^. Additional pathways that remodel the inflammatory phenotype and promote tumor growth or proliferation, and were associated with higher risk, included alpha protein kinase 1 ^45^, TRAF6-mediated NFKB activation ^46^, and oncogene-induced senescence pathways ^47^.

## 3 Discussion

In this study, we undertook a comprehensive radiogenomic analysis of pLGGs using both supervised and unsupervised ML techniques. Our primary objective is to pave the path for curing pLGGs while reducing the need for invasive tumor resection or systemic treatments, thereby reducing related morbidities in children. We focused on identifying intrinsic imaging and molecular characteristics of pLGG tumors, exploring their phenotype-genotype relationships from various analytical perspectives for distinct clinical applications. This method also addresses a major hurdle in ML: the lack of biological or clinical interpretability which hinders its integration into treatment decisions.

The analysis of immunological clusters revealed their association with clinical outcomes, wherein immune-high cluster 2 was associated with worst progression-free survival rates, likely linked to enrichment of M2-polarized macrophages, which suppress local tumor immunity and mast cells reshaping the TIME to promote angiogenesis ^48,49^. Conversely, memory B-cells that favor anti-tumor T-cell mediated immunity in glioma ^36^ were relatively low in cluster 2 and higher in prognostically-favorable cluster 1.

Immune cluster 1, with more favorable prognosis, showed relatively higher inferred levels of eosinophils, also observed in independent LGG cohorts ^50^. In cluster 3, which showed the longest median overall survival of all immune groups, we detected signatures related to NK and CD8+ cell types, both reported to confer more favorable prognosis in glioblastoma ^51–54^. This suggests that profiling immune cell types in pLGG may provide insights with respect to disease prognosis.

These findings also suggest the potential benefit of immune clustering in determining response to and stratifying patients for checkpoint inhibitor therapy. *BRAF* is vital in the *MAPK* signaling pathway, influencing various cellular functions like proliferation and angiogenesis ^55^. Although *BRAF* inhibitors are potential treatments in pLGGs ^56,57^, there is a prevalent issue of therapeutic resistance ^58^ and serious side effects like secondary neoplasms ^59^. As such, exploring alternative treatments like immunotherapy, either on its own or combined with targeted therapies, may be beneficial for pLGGs with *BRAF* mutations ^60^. In our study, we discovered that *BRAF*-altered pLGGs exhibit increased immune cell infiltration-related gene expression. It has been shown that the *BRAF V600E* mutation may boost immunogenicity in LGG patients, leading to the development of *BRAF V600E*-specific T-cells, which can then be targeted with T-cell therapy ^61^. However, another study focusing on pLGGs found no link between PD-1 expression levels and the *BRAF V600E* mutation status ^62^. As we continue to make advancements in molecular research, further investigations will be essential to pinpoint the optimal immunotherapeutic strategies for pLGG patients.

In our research, a strong correlation was found between higher TIS and immunological groups in pLGGs, whereas TMB was relatively high in only one cluster with relatively inferred immune cell content (Cluster 1). This is consistent with a previous study on angiosarcoma patients where TIS was linked to specific immune clusters, including neutrophils, macrophages, and PD-L1+ cells ^63^. Furthermore, a breast cancer study found TIS scores closely aligned with tumor-infiltrating lymphocyte numbers ^64^. Pan-cancer studies have also recognized TIS as a consistent predictor of immunotherapy response across multiple cancers ^17^.

Wang et al. ^65^ previously reported the immunological classification of pediatric gliomas, including both low- and high-grade tumors, based on RNA-sequencing. They identified three primary immunological subgroups, with the immune-hot group showing a higher presence of M2-polarized macrophages and CD4+ Memory T-cells, which aligns with our findings. However, unlike their study, we did not observe any differences in OS among the three immunological subgroups. This discrepancy could primarily be due to Wang et al.’s inclusion of high-grade tumors in their analysis. In contrast, pLGGs usually have a prolonged OS, leading to a higher proportion of censored data in our study. In our research, we advanced the methodology by developing an imaging-based radioimmunomic signature that predicts immunological clusters. This signature acts as a surrogate biomarker for immunological profiles in situations where RNA-sequencing is unfeasible, such as when the tumor location impedes surgical resection, or in settings where RNA-sequencing and transcriptomic analysis are not possible due to limited resources.

The ability of our imaging-based ML model to distinguish the ‘immune-hot’ cluster 2 from immune clusters 1 and 3, each harboring distinct cellular profiles, may prove clinically beneficial when it comes to predicting the efficacy of immune-based therapies in pLGG upfront. Radiomics has been used for predicting immune phenotypes ^66^ and specific immune cell populations ^67–69^ in adult glioblastoma. Nonetheless, our investigation stands as the pioneering effort to create imaging-based ML models capable of predicting immune clusters in pediatric brain tumors. Our study highlights the value of integrating ADC-derived features that provide quantitative assessments of pattern of diffusivity of water molecules in the TIME. The reports on the positive or negative correlation of ADC values and tumor microenvironment markers remain contradictory ^68–70^. Moreover, a radiomic study based solely on ADC maps did not outperform a model constructed using post-contrast T1-weighted MRI sequences, indicating the complexity of identifying the most informative imaging features for such predictive tasks ^66^. While studies in the adult brain tumor domain have pointed to the improved accuracy of radiogenomic classifications after the inclusion of features from advanced MRI sequences such as ADC map ^8,71^, there remains a paucity of research that directly correlates ADC values with the TIME in brain tumors ^69,70^.

We sought to determine the predictive value of clinically significant variables, known for their prognostic impact in pLGG, and radiomic features from pre-surgical MRIs for assessing tumor progression risk. The high reproducibility of our clinicoradiomic model, evident with concordance indexes of 0.71 and 0.77 in the discovery and replication sets, respectively, indicates its potential for future application in unseen patient cohorts, including prospective studies. Consistent with existing literature ^72,73^, the extent of tumor resection was a key determinant of progression-free survival. In our study, among 61 progressed tumors, only 7 had undergone total or near-total resection, with progression-free intervals ranging from 222 to 921 days and a median of 660 days post-surgery.

When comparing our clinicoradiomic risk groups with treatment risk categories, we found that most treatment high-risk tumors were also identified as clinicoradiomic high-risk tumors, while a few were in the medium-risk category. Most treatment low-risk tumors corresponded to our low to medium-risk clinicoradiomic groups. Notably, the treatment low-risk tumors categorized as high-risk by our model still progressed within five years, (progression-free survival range 4.8 - 131.5, median, 19.2 months), suggesting accuracy of our model in predicting progression-free survival and progression risk. These findings underscore our model’s potential to guide treatment strategies, suggesting consideration for a targeted therapy approach or enrollment onto a clinical trial upfront, rather than at recurrence, for ‘high-risk’ tumors and more conservative, monitoring-focused approaches for ‘low-risk’ cases. The ‘medium-risk’ category could be considered for a proactive treatment approach ^3^, although it currently lacks a clear clinical definition in pLGG tumors, which complicates understanding its clinical implications. However, our findings indicate an over-representation of *SYNE1* gene, linked to cerebellar ataxia and tumor progression, in the medium-risk clinicoradiomic group. Considering the long-term morbidities associated with pLGG tumors, including ataxia, children in this risk category may be predisposed to the development of progressive ataxia ^74^, underscoring the importance of upfront risk stratification to either prevent the onset or enable early management of these side effects.

Biological interpretation of our clinicoradiomic model via radiogenomic analysis showed that the predicted risk scores were associated with molecular pathways known to influence tumor progression which may provide insights for clinical interventions. *NFKB* signaling, a pathway known for the maintenance of B-cells and its role in T-cell driven anti-tumor immune responses in glioblastoma ^45–47^, was negatively associated with risk. Pathways related to the balance of fatty acid synthesis and oxidation, as a metabolic switch to promote lipogenesis and lipid droplet formation – a well-established tumor-promoting mechanism across cancers particularly high-grade gliomas ^41,42^ – were found to relate to elevated risk. IL-3, which suppresses anti-tumor immune responses via activation of the PDL1 pathway ^42^, and IL-5 signaling, shown to promote cancer metastasis by remodeling the immune microenvironment ^43^, were also linked to higher risk.

The existing literature on radiomic analysis for pediatric brain low-grade glioma is limited, primarily concentrating on the prediction of single genetic mutations or alterations, such as *BRAF-V600E* mutation and *BRAF-KIAA1549* fusion ^9–12^. In light of the 2021 WHO classification of tumors of the central nervous system (CNS), 5^th^ edition (WHO CNS 5) ^2^, which recommends the inclusion of driver molecular alterations in standard diagnostic processes, merely predicting a single mutation or fusion using imaging might not yield substantial clinical benefits. Our study is the first to offer a comprehensive radiogenomic analysis that integrates radiomics with transcriptomic pathways, extending beyond the predictive modeling of single molecular drivers and addressing a research gap in pediatric and adult brain tumor studies.

The current study encountered challenges due to the limited availability of imaging and genomic data (WGS or RNA-sequencing), compounded by the rarity of pediatric brain tumors. The cohort comprised a limited number of patients with *NF1* disease, non-pilocytic astrocytoma histologies, and was confined to specific tumor sites. The time of progression was determined by reviewing clinical notes on treatment changes and MRI scans when progression was suspected, though this method may not be entirely objective. Future studies could improve accuracy by employing volumetric analysis, although its advantage over the traditional bidimensional method is not yet proven. Our analysis comprised radiomic, genomic, and transcriptomic data, whereas other important layers like methylation profiling, metabolomics, proteomics, and pathomics were not available for most of the patients. To gain a deeper understanding of the complex interplay between these data layers at micro and macro scales, and how they influence the clinical presentation and treatment response of tumors, future studies incorporating a broader spectrum of data could further inform precision medicine approaches for individual patients. Additionally, due to unavailable tissue, we were unable to validate our transcriptomic findings through immunohistochemical staining, suggesting a need for further research to confirm cell enrichments in immunological subgroups.

Acknowledging these limitations, our analysis sets the stage for the development of more personalized and less invasive treatment strategies for pLGGs in future research. Specifically:

- The discovery of the radioimmunomic signature provides a non-invasive technique to identify immune-high pLGG tumors, with elevated TIS levels and typically a poorer prognosis, which may be candidates for immunotherapy. This approach can aid in the early stratification of patients, allowing for their inclusion in immunotherapy trials.
- The clinicoradiomic predictive model can serve as an effective marker for assessing patient’s risk of progression and predicting treatment response. This model can be applied as a pre-interventional assessment tool, enabling consideration of different scenarios, such as varying degrees of surgical resection and chemotherapy use. Its predictive capability allows for the simulation of diverse clinical outcomes based on hypothetical treatment strategies prior to making patient management decisions. Moreover, the predictions of the clinicoradiomic model can be updated and recalibrated after each intervention, or as new data about tumor molecular subtypes emerge post-resection. This iterative refinement ensures that the prognostic evaluations stay accurate and applicable to the patient’s changing clinical condition. Future enhancements to the model, including the incorporation of multi-omic data, will further refine its accuracy and utility. Integrating this predictive model into clinical practice will provide valuable guidance in treatment planning, thereby enhancing the management and care of patients with pLGGs.

Overall, by leveraging the insights from our analysis, a more patient-centric approach can be adopted, tailoring treatments to the specific characteristics and needs of each patient, thus enhancing the overall quality and effectiveness of care for patients with pLGGs, aligning closely with the evolving field of personalized medicine.

## 4 Online Methods

### 4.1 Data

#### 4.1.1 Overview of the Data

In this HIPAA-compliant study, data from patients diagnosed histopathologically with *de novo* pediatric low-grade glioma (pLGG), retrospectively collected through the Children’s Brain Tumor Network (CBTN) ^13^, was included. RNA-sequencing data was available for 494 subjects from a combination of CBTN patient data and patient data available in the OpenPedCan repository (v12 data release), which is a collection of genomic and molecular data available on multiple sources, such as KidsFirst ^75^, TARGET ^76^, OpenPBTA ^77,78^, and the TCGA ^79^. Subjects had molecular subtype information available through the OpenPedCan database and through the application of an in-house molecular subtyping pipeline on additional cases consented on CBTN’s research protocol after the v12 OpenPedCan release ^77^.

Treatment-naïve multiparametric MRI (mpMRI) sequences, including pre- and post-Gadolinium T1-weighted (T1w, T1w-Gd), T2-weighted (T2w), and T2 fluid attenuated inversion recovery (T2-FLAIR), acquired between 2006 and 2018, from 258 subjects with pLGG were obtained. Diffusion weighted imaging (DWI) and scanner-generated apparent diffusion coefficient (ADC) maps were collected for 153 subjects. Patients were excluded if: (1) age at the time of imaging was older than 18 years, (2) the images were not treatment-naïve, i.e., were not acquired prior to the initial surgery or treatment, (3) the primary site was not brain, (4) leptomeningeal dissemination was reported, (5) not all four standard mpMRI sequences were available, or (6) the images were determined to be of low-quality. A final cohort of 201 subjects with available standard mpMRI (91 with ADC-maps) was included in the imaging studies. In cases where multiple imaging sessions were available for the selected patients before their initial treatment or surgery, we chose the session that was closest in time to the intervention. The time interval between these sessions and the intervention for our cohort ranged from 1 to 370 days, with a median interval of 3 days.

For the patients in the final cohort, we reviewed the clinical notes for the patients that had progressed. For the subjects that had tumor progression, we recorded the number of radiotherapy or chemotherapy treatments the patients had received over the course of the study within the five years since the initial diagnosis. A treatment change was classified as a new treatment only in cases of treatment failure, not when a change in treatment method was required due to toxicity.

#### 4.1.2 Molecular Data Analysis

4.1.2.1 *Enrichment analysis and clustering of immune and stromal cell types*

To identify the enrichment of immune and stromal cell types and pathways, xCell ^18^, which is a cell type enrichment analysis from gene expression data, was applied on our pLGG transcriptomic data. We leveraged transcripts per million (TPM) generated by a harmonized STAR-RSEM RNA-sequencing pipeline developed as part of the Gabriella Miller Kids First Data Resource Center sequencing and benchmarking effort ^78^, which newly leverages Ensembl’s GENCODE version 39 ^80^ for gene-level annotations. We leveraged the Immunedeconv R package ^81^ specifying default parameters and xCell as the method of interest. Tumor purity was inferred using three different approaches, including estimates of tumor purity derived from copy number (ABSOLUTE) ^14^, expression (ESTIMATE) ^15^, and methylation-based assessments (LUMP) ^16^ (Supplemental Figure 1). Kruskal-Wallis’s test was performed to determine the differences in tumor inflammation signature (TIS) and tumor mutational burden (TMB) levels among the immune clusters.

##### 4.1.2.2 Enrichment analysis of molecular pathways

To identify differentially expressed biological pathways across clinicoradiomic risk groups, we filtered gene expression data for the ‘protein coding’ gene type. We then provided expected counts as input to DeSeq2 ^82^ for differential gene expression analysis, specifying clinicoradiomic risk groups as the factor variable in the model’s design. Lists of differentially expressed genes derived from pairwise comparisons across all three imaging groups were ranked and used as input for pre-ranked GSEA ^83^ to identify differentially expressed biological pathways. Pathway annotations were derived from Reactome using the Molecular Signatures Database v2023.1.Hs ^84^.

##### 4.1.2.3 Germline Variant Analysis

To identify deleterious/likely deleterious germline variants in a list of cancer predisposition genes, we applied the following criteria to filter out single nucleotide variants (SNVs) and small insertions/deletions (indels): a) minor allele frequency in gnomAD (v2.1.1 ^85^) and TOPMed (data freeze 7 ^86^) is less than or equal to 0.0001; b) at least half of 21 prediction scores as listed in Supplementary Table X of the dbNSFP database (v4.2 ^87,88^) label a variant as Deleterious, Detrimental, Damaging or Medium/High Impact; c) for quality control purposes we require that in heterozygous calls, alternate allele depth is between 25% and 75% of all sequencing depth of a variant site. We also checked if variants are cataloged in either ClinVar (release 20240331 ^89^) or Human Gene Mutation Database (HGMD; Professional Version 2024Q1 ^90^). Variants with membership in ClinVar/HGMD are considered “deleterious”, while others obtained through the germline variant filtering process described above are considered “likely deleterious”.

#### 4.1.3 Imaging Data Processing and Analysis

In our study, image standardization was performed prior to radiomic feature extraction. For each patient, all MRI scans were first re-oriented to left-posterior-superior (LPS) coordinate system. Subsequently, T1w, T2w, T2-FLAIR, and ADC-maps (when available) were co-registered with their corresponding T1w-Gd sequence. All images were then resampled to an isotropic resolution of 1 *mm*3 based on anatomical SRI24 atlas using the Greedy algorithm, in Cancer Imaging Phenomics Toolkit open-source software v.1.8.1 (CaPTk, https://www.cbica.upenn.edu/captk). All preprocessed images were then skull-stripped using our dedicated brain tissue extraction tool ^91^. The skull-stripped images were then normalized to the intensity range of [0, 255] after removing after removing the outlier pixels that did not fall into 99.9% percentile of the image histogram. Brain tumor segmentation was performed using our in-house automatic pediatric brain tumor segmentation tool, described in ^91^, and manual revisions were made when necessary. This tool generates segmentation of tumor subregions, including the enhancing tumor, nonenhancing tumor, cyst, and edema. A combination of all subregions is used to generate a whole lesion segmentation (indicated in Figure 1).

Radiomic features (n = 881) of shape, volume, intensity, first-order histogram (10 bins), gray-level co-occurrence matrix (GLCM), gray-level run-length matrix (GLRLM), gray-level size zone matrix (GLSZM), neighborhood gray tone difference matrix (NGTDM), local binary pattern (LBP), and Collage features were extracted after overlaying the WT mask on the mpMR images.

The data was randomly split into a “discovery” (n = 160) and an unseen “replication” (n = 41) cohort; since some tumor locations, such as basal ganglia, were rare in our cohort, we kept those subjects in the discovery set. We have used the same cohort split across all experiments throughout this study; however, some of the experiments had reduced sample sizes when intersected with transcriptomic data. Prior to each ML analyses, pairwise Pearson’s correlation was performed on the extracted radiomic features for the subjects in the discovery cohort, to remove one of the highly correlated pair of features (*r* > 0.90), reducing the dimension to 438. The features were then normalized using z-scoring approach. For categorical variables, including sex, *NF1* disease, tumor location, extent of tumor resection, chemotherapy, and radiotherapy, count encoding was performed. Age at diagnosis, as a continuous variable, was normalized using z-scoring method. The mean and standard deviation calculated for the continuous features in the discovery set were used to normalize the corresponding features in the replication set.

Histograms illustrating the most indicative features were produced for different groups of tumors in each of the experiments, to elucidate the inherent biological processes associated with specific genetic alterations. These histograms display the frequency (y-axis) of a particular feature value (x-axis), collated from the entire patient dataset.

### 4.2 Imaging-Based Prediction of TIME Subgroups via Radioimmunomic Signature

For this analysis, immunological subgroups of pLGG were determined using unsupervised clustering based on enrichment of immune and stromal cells (obtained from xCell), followed by application of supervised approach based on radiomic features for classification of the immune clusters (Figure 1 shows the workflow for this analysis).

#### 4.2.1 Characterization of TIME Subgroups

We leveraged the xCell enrichment scores derived as described above as input for clustering. Clustering was carried out using an internally developed R package that comparatively and quantitatively assesses clustering quality across dissimilarity-based ^92^, model-based ^93^, joint dimensionality reduction-based ^94^, and dbscan ^95^ clustering methods. We leveraged the fpc R package to derive a quantitative composite score using multiple metrics of clustering quality, selecting the highest scoring method based on cluster compactness and separation. The v-test, which is a measure of the difference in means across a given cluster relative to the remaining sample set scaled by the variance ^96^, was leveraged for identifying cluster-specific immune cell subsets. Heatmap plots were generated using the pheatmap R package, violin plots using ggplot2 R package, and radar plots using the fmsb R package.

#### 4.2.2 Radioimmunomic Signature

To generate a radiomic signature of immunological profiles, we used the intersection of our imaging and transcriptomic cohorts, resulting in a total of 150 subjects (120 in the discovery, 30 in the replication sets) with all four conventional MRI sequences (T1w, T1w-Gd, T2w, T2-FLAIR) and 91 subjects (72 in the discovery, 19 in the replication sets) when ADC-map was also available. On a feature set including the radiomic features (n = 438 when the four conventional MRI sequences were available and n = 493 when ADC was also included) and age at diagnosis, support vector machines (SVM), wrapped with Maximum Relevance Minimum Redundancy (MRMR) feature selection, was applied to classify the subjects in the discovery set into three immunological groups. ML classifier was trained using a nested cross-validation (CV) approach, with five folds for the inner loop for the purpose of feature selection and ten folds for the outer loop to tune the hyperparameters and optimize the model. This nested CV approach is intended to avoid data overfitting and improve generalizability to the unseen replication cohort. To reduce overfitting to the training data, we used a linear kernel for SVM. Three binary classifiers for each of the immunological groups were trained in a one-versus-the-rest approach. This implementation was carried out in MATLAB 2023a (MathWorks Inc). The generated ML models were then independently applied to the replication set to evaluate the performance via area under the receiver operating characteristic (ROC) curve (AUC) and balanced accuracy.

### 4.3 Imaging-Based Prediction of Risk of Tumor Progression

Patient demographics and clinical variables considered to play an important role in determining the risk of the patient for tumor progression, i.e., age, sex, extent of tumor resection, treatment method (radiotherapy, chemotherapy), tumor location, and germline Neurofibromatosis (*NF1*) cancer predisposition were included as input variables. A baseline Cox-PH model only based on clinical variables was generated to study additional value of radiomics to these clinically available features. The data (n = 201) was split into a discovery (n = 160) and replication (n = 41) sets. The discovery set was used for training a Cox-PH model using a stratified nested cross-validation (10 folds for both inner and outer nests) approach. The trained model was applied on the “unseen” replication set. Consequently, we generated another Cox-PH model with ElasticNet penalization using a combination of clinical and radiomic variables. The penalization was set to zero for clinical variables but applied for radiomic features to decrease the dimensionality of the feature space. The performances of survival models were evaluated based on three metrics were employed: 1) Harrell’s concordance index (hereinafter referred to as “HAR”), 2) Uno’s concordance index (hereinafter referred to as “UNO”), and 3) the Integrated Brier Score (hereinafter referred to as “IBS”), which provides an overall calculation of the model performance at all available times. The patients were then stratified into the groups of low, medium, and high risk based on their predicted risk in the discovery set. To achieve this, we fitted the entire discovery cohort against the survival model to obtain risk scores. Based on the distribution of risk scores calculated by the model, the patients were grouped into three categories: 1) High (>=75%), 2) Medium (>=25%; <75%), and 3) Low (< 25%).

### 4.4 Statistical Analysis

Tumor mutational burden (TMB) and tumor inflammation signature (TIS) were calculated from the whole genome sequencing (WGS) data using the R package “maftools” based on VarScan method. Molecular Signatures Database (MSigDB, http://www.gsea-msigdb.org/gsea/index.jsp) was used to extract immune or inflammation gene sets and differential expression analysis was performed between tumor and normal tissues. Analysis of correlation between the categorical variables from the discovered immune groups and pLGG molecular subtypes was carried out using chi-squared test. Kruskal-Wallis and one-way analysis of variance (ANOVA) tests were performed to evaluate the statistical difference in the mean values of TMB and TIS, respectively, across the three different immune groups. Student’s t-test was applied to calculate the difference between the mean values of the groups of patients that had progressed and had no or one treatment and more than one treatment. For evaluation of the performance of the multivariate classification approach, area under the receiver operating characteristic (ROC) curve (AUC) along with 95% confidence interval (95% CI) were calculated.

### 4.5 Data and Code Availability Statements

#### 4.5.1 Data Availability

De-identified genomic and transcriptomic source data as well as clinical data, including patient baseline characteristics, molecular subtypes, and outcomes, can be found under the dbGaP study phs002517.v2.p2 as well as through the Kids First Data Resource Portal. The imaging data is available through Children’s Brain Tumor Network (CBTN, cbtn.org). The processed data is available at https://github.com/d3b-center/pLGG-immune-clinicoradiomics.

#### 4.5.2 Code Availability

The code for analyzing processed clinical, genomic, transcriptomic, and radiomic data in relation to patient clinical characteristics can be found at https://github.com/d3b-center/pLGG-immune-clinicoradiomics. All image processing tools used in this study are freely available for public use (CaPTk, https://www.cbica.upenn.edu/captk; ITK-SNAP, https://www.itksnap.org; the in-house automated tumor segmentation model, https://github.com/d3b-center/peds-brain-auto-seg-public).

## Data Availability

All data produced in the present study are available to the public (see Data Availability Statement)

https://github.com/d3b-center/pLGG-immune-clinicoradiomics

https://doi.org/10.5281/zenodo.13942516

