## Supplementary Tables and Figures for "Multiparametric MRI Along with Machine Learning Informs on Molecular Underpinnings, Prognosis, and Treatment Response in Pediatric Low-Grade Glioma"

### 6 Supplementary Information

#### 6.1 Imaging Data Description

Supplemental Table 1. Summary of demographics and clinical characteristics of the pLGG patients included in the imaging cohort (collected through the CBTN data repository)

| Variable | Value | # of Patients |
| --- | --- | --- |
| Age Range (months) | 4.3 – 280.73; Mean, 108.07 | 201 |
| Sex | Female | 97 |
|  | Male | 104 |
|  | Not Reported |  |
| Tumor Location | Basal Ganglia | 2 |
|  | Brainstem | 10 |
|  | Cerebellar | 86 |
|  | Intraventricular | 5 |
|  | Lobar (frontal, parietal, temporal, or occipital lobes) | 44 |
|  | Multifocal | 14 |
|  | Suprasellar | 34 |
|  | Thalamus | 6 |
| NF1 Disease | With *NF1* disease | 8 |
|  | Without *NF1* disease | 193 |
| Extent of Tumor Resection | Gross or near total resection | 109 |
|  | Partial resection | 58 |
|  | Biopsy | 27 |
|  | Not Reported/Unavailable | 7 |
| Treatment | Radiotherapy | 10 |
|  | Chemotherapy | 46 |
|  | None or Not Available | 149 |
| Molecular Subtypes | LGG, *KIAA1549-BRAF* | 68 |
|  | LGG, *BRAF V600E* | 14 |
|  | LGG, *CDKN2A/B* | 4 |
|  | LGG, *wildtype* | 19 |
|  | LGG, *IDH* | 1 |
|  | LGG, *NF1* | 8 |
|  | LGG, *RTK* | 11 |
|  | LGG, other *MAPK* | 11 |
|  | LGG, *MYB/MYBL1* | 1 |
|  | GNG, *KIAA1549-BRAF* | 3 |
|  | GNG, *wildtype* | 5 |
|  | Other LGG or GNG | 5 |
|  | Not Available | 51 |
| Progression-Free Survival Range (months) | 2.83 – 133.37  Mean, 39.88 | 201 |
| Number of Treatments for Progressed Tumors | Zero or one | 31 |
|  | More than one treatment | 30 |

#### 6.2 XCell Immunological Clusters

We observed a significant negative correlation between immune score and estimates of tumor purity derived from copy number (ABSOLUTE) [87], expression (ESTIMATE) [88], and methylation-based assessments (LUMP) [89] (Supplementary Figure 1).

| 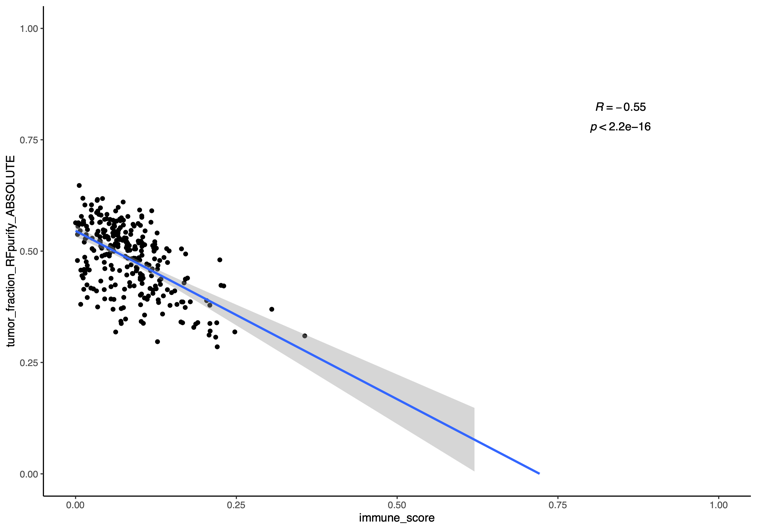 | 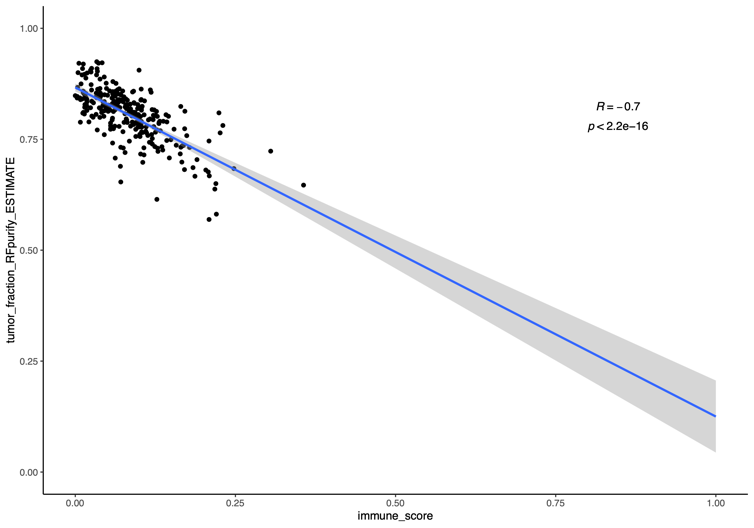 |
| --- | --- |
| 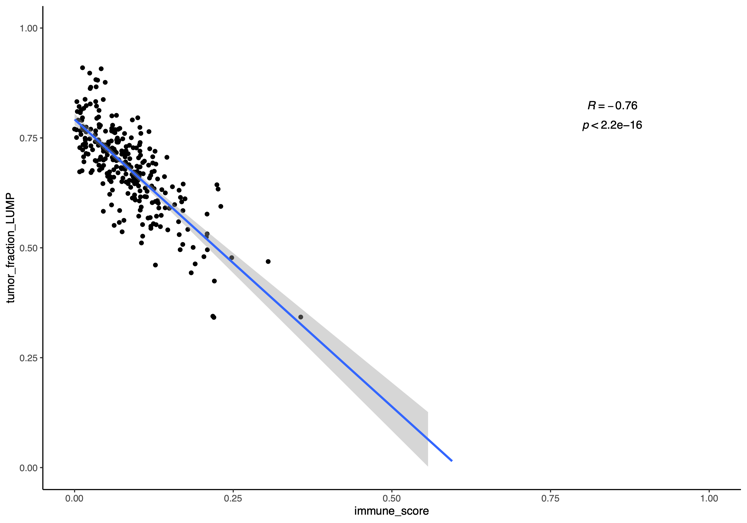 |  |

Supplemental Figure 1. Tumor purity obtained using ABSOLUTE, ESTIMATE, and LUMP, plotted against immune score.

Supplemental Table 2. The Cox effect results for the analysis of progression-free survival

|  | coef | exp(coef) | se(coef) | z | Pr(>\|z\|) |
| --- | --- | --- | --- | --- | --- |
| age_at_diagnosis_days | -0.0001129 | 0.99988714 | 3.02E-05 | -3.7424565 | 0.00018223 |
| reported_genderMale | 0.01036621 | 1.01042012 | 0.11006651 | 0.09418132 | 0.92496512 |
| raceAsian | 0.46689584 | 1.59503526 | 0.70135233 | 0.66570798 | 0.50559777 |
| raceBlack or African American | 0.34875673 | 1.41730436 | 0.63098567 | 0.55271736 | 0.58045697 |
| raceMore Than One Race | -0.3151572 | 0.72967416 | 0.85511459 | -0.3685555 | 0.71245906 |
| raceNative Hawaiian or Other Pacific Islander | 4.02664876 | 56.0726828 | 1.23658708 | 3.25625977 | 0.0011289 |
| raceReported Unknown | -0.2903477 | 0.74800341 | 0.62580974 | -0.4639553 | 0.64267977 |
| raceWhite | -0.0591466 | 0.9425686 | 0.61640544 | -0.095954 | 0.9235571 |
| CNS_regionMidline | 0.28999767 | 1.33642438 | 0.2413468 | 1.20158078 | 0.22952599 |
| CNS_regionMixed | 0.40111046 | 1.49348223 | 0.19765785 | 2.02931713 | 0.042426 |
| CNS_regionOptic pathway | -0.0865064 | 0.91712964 | 0.44250286 | -0.1954935 | 0.84500662 |
| CNS_regionOther | 0.33205425 | 1.39382846 | 0.57129883 | 0.5812269 | 0.56108754 |
| CNS_regionPosterior fossa | 0.11129102 | 1.11772014 | 0.16613339 | 0.66988954 | 0.50292821 |
| CNS_regionSpine | -0.1032594 | 0.90189296 | 0.37048277 | -0.2787159 | 0.78046285 |
| CNS_regionSuprasellar | 0.43204445 | 1.54040359 | 0.32432297 | 1.33214262 | 0.18281333 |
| CNS_regionVentricles | 0.44623032 | 1.56241128 | 0.32673308 | 1.36573352 | 0.17202262 |
| molecular_subtypeGNG, MYB/MYBL1 | 1.37374301 | 3.95010835 | 0.81972593 | 1.67585648 | 0.09376631 |
| molecular_subtypeGNG, other MAPK | 0.76562168 | 2.15033077 | 0.64556099 | 1.18597884 | 0.23563066 |
| molecular_subtypeGNG, to be classified | 0.08704222 | 1.09094274 | 0.84451776 | 0.10306737 | 0.91790951 |
| molecular_subtypeGNG, wildtype | 0.53513131 | 1.70767246 | 0.45890701 | 1.16609967 | 0.24357415 |
| molecular_subtypeGNT, KIAA1549-BRAF | 0.63261592 | 1.88252869 | 1.0861798 | 0.58242284 | 0.5602819 |
| molecular_subtypeGNT, other MAPK | 0.00184952 | 1.00185123 | 1.13558941 | 0.00162869 | 0.9987005 |
| molecular_subtypeGNT, wildtype | 1.05593068 | 2.87464928 | 1.08180781 | 0.97607973 | 0.32902495 |
| molecular_subtypeLGG_CDKN2A/B | 0.59495645 | 1.81295198 | 0.48489504 | 1.22697985 | 0.21983017 |
| molecular_subtypeLGG_IDH | 0.67410583 | 1.96227758 | 0.5504056 | 1.22474377 | 0.22067178 |
| molecular_subtypeLGG_NF1 | 0.09630276 | 1.10109238 | 0.4578858 | 0.21032048 | 0.83341755 |
| molecular_subtypeLGG_RTK | 0.35382055 | 1.42449954 | 0.41504 | 0.85249747 | 0.39393804 |
| molecular_subtypeLGG, BRAF V600E | 0.43084801 | 1.53856168 | 0.42088349 | 1.02367523 | 0.3059887 |
| molecular_subtypeLGG, H3, other MAPK | 0.19335371 | 1.21331188 | 1.08547977 | 0.17812742 | 0.85862291 |
| molecular_subtypeLGG, KIAA1549-BRAF | 0.30041245 | 1.35041567 | 0.4023964 | 0.74655849 | 0.45533011 |
| molecular_subtypeLGG, KIAA1549-BRAF, other MAPK | 1.4846357 | 4.41335735 | 0.70614106 | 2.10246336 | 0.03551271 |
| molecular_subtypeLGG, MYB/MYBL1 | 0.38898619 | 1.47548417 | 0.63249974 | 0.61499818 | 0.53855592 |
| molecular_subtypeLGG, other MAPK | 0.89592262 | 2.4495948 | 0.44944005 | 1.99341963 | 0.04621552 |
| molecular_subtypeLGG, To be classified | -0.6311619 | 0.53197337 | 1.08667639 | -0.5808186 | 0.56136272 |
| molecular_subtypeLGG, wildtype | 0.53212698 | 1.70254974 | 0.40427322 | 1.31625582 | 0.18808819 |
| molecular_subtypeSEGA, wildtype | -0.4663222 | 0.62730511 | 0.57104384 | -0.8166137 | 0.4141492 |
| clusterID2 | 0.49963731 | 1.64812341 | 0.13462076 | 3.71144314 | 0.00020608 |
| clusterID3 | 0.10565895 | 1.11144276 | 0.1511455 | 0.69905456 | 0.48451793 |

| 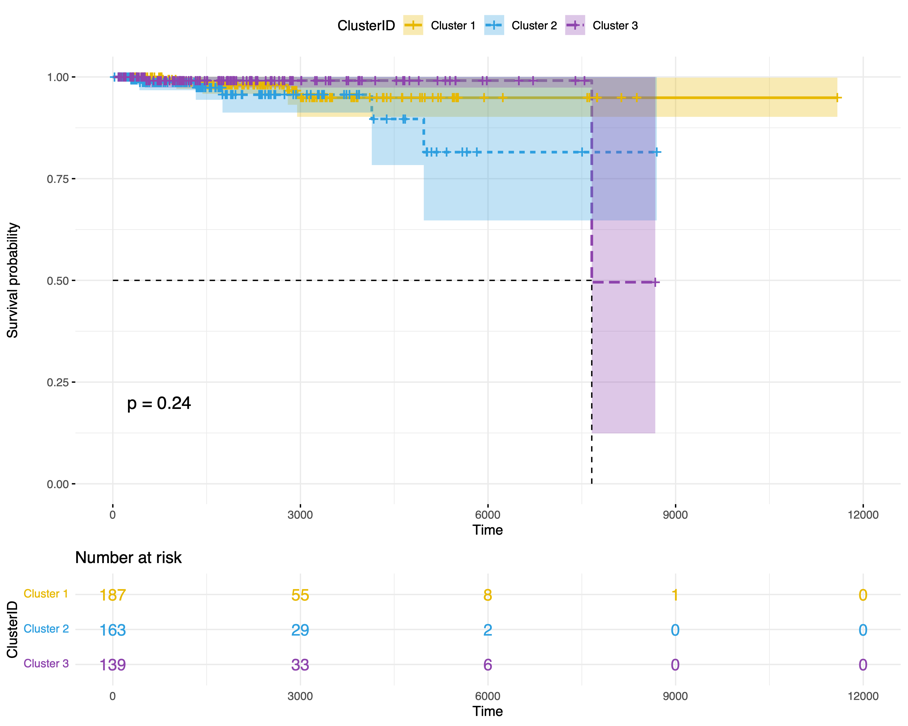 | 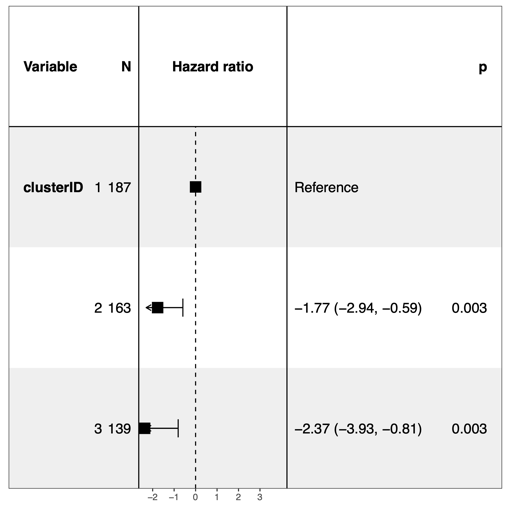 |
| --- | --- |

Supplemental Figure 2. Kaplan-Meier curves presenting the overall survival probability for patients in the three xCell clusters along with the forest plot.

| 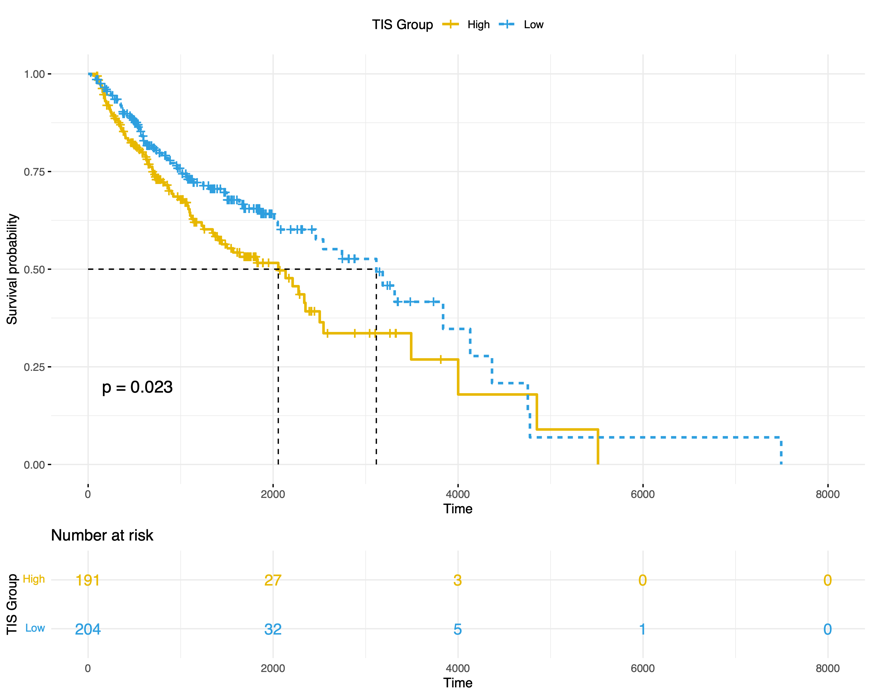  **(A)** | 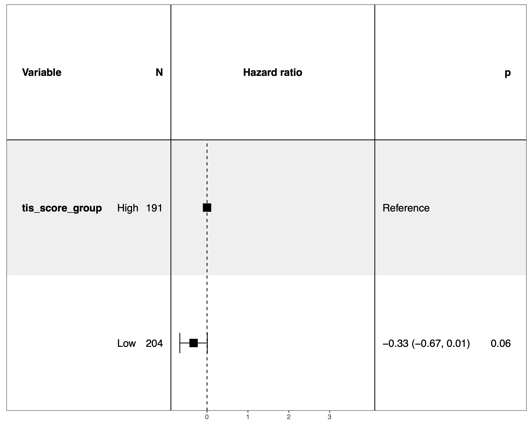  **(B)** |
| --- | --- |
| 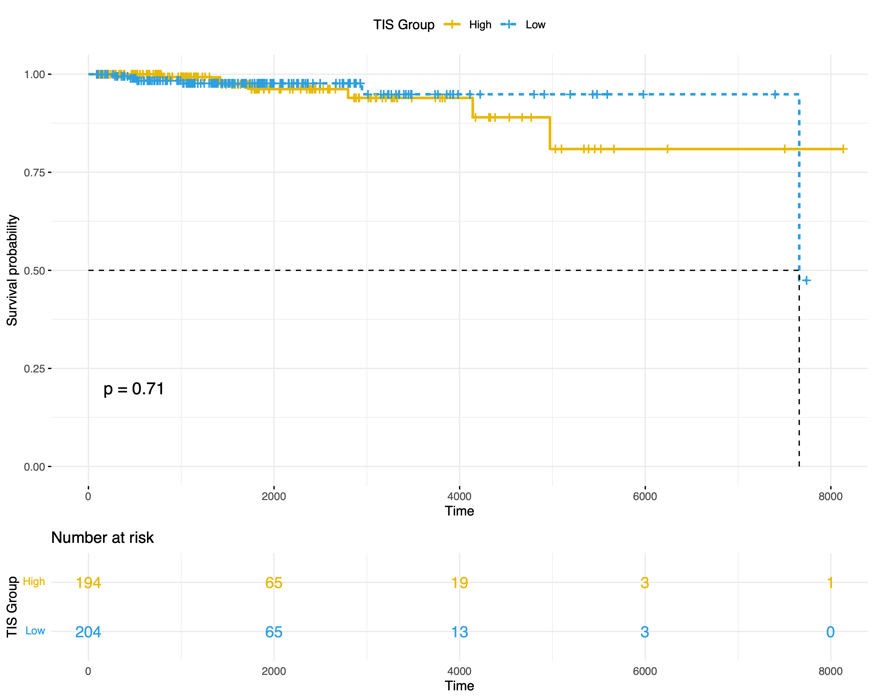  **(C)** | 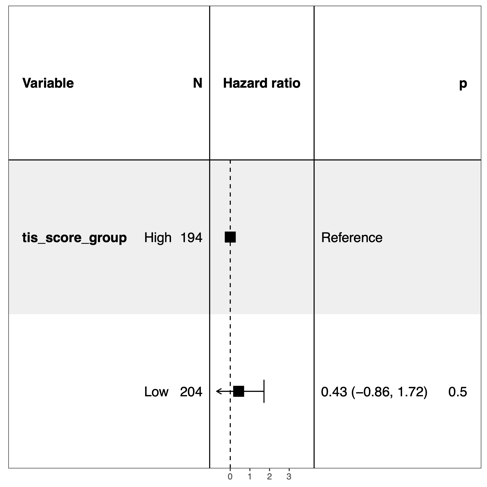  **(D)** |

Supplemental Figure 3. Kaplan-Meier curves indicating progression-free survival probability (Top Row) and overall survival (Bottom Row) for the patients with high versus low levels of Tumor Inflammation Signature (TIS), along with the associated forest plots.

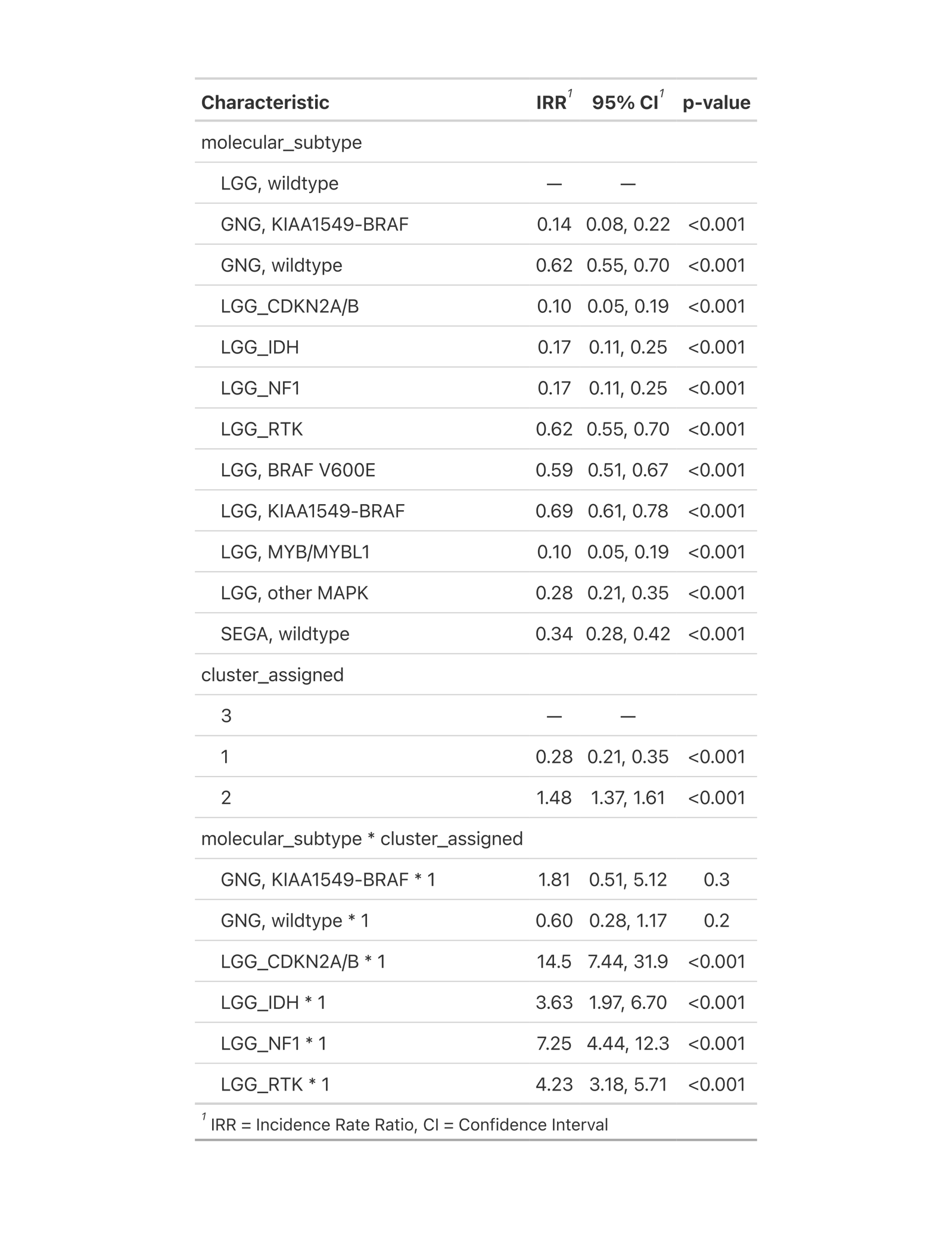

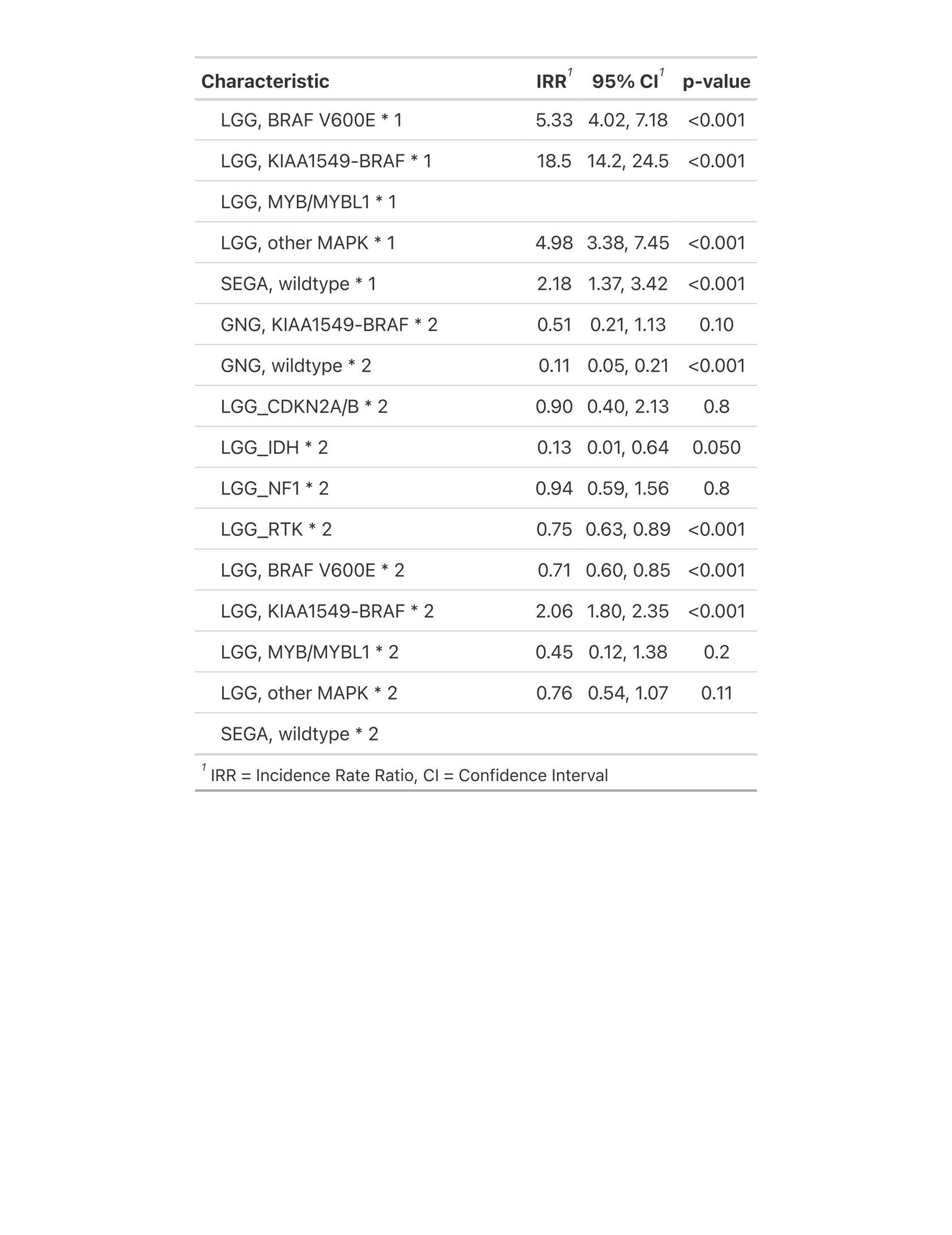

Supplemental Figure 4. Summary of Poisson generalized linear model parameters comparing counts across pLGG XCell clusters and molecular subtypes.

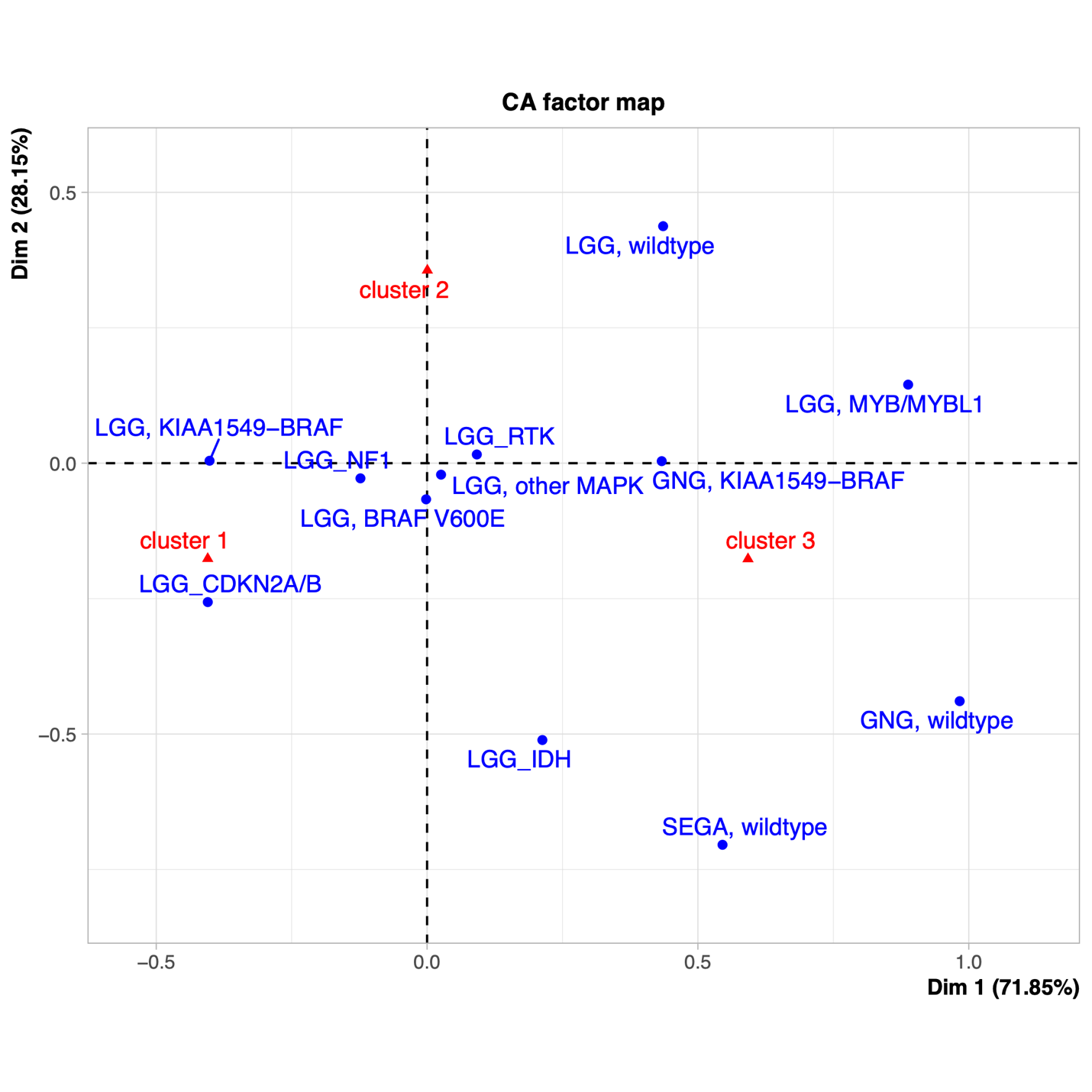

Supplemental Figure 5. Correspondence analysis of Pearson residuals from Poisson generalized linear model showing the association between pLGG molecular subtypes and XCell clusters.

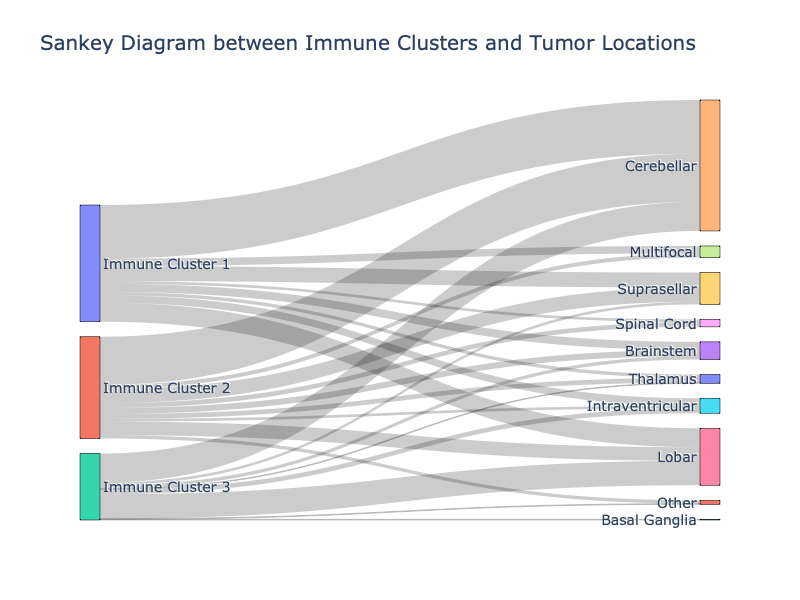

Supplemental Figure 6. Sankey plot of the association of imaging clusters (described in Section 2.2), immunological clusters, and tumor locations.

#### 6.3 Risk Stratification

##### 6.3.1 Model 1: Clinical-Only Variables

We first investigated the performance of a Cox-PH model trained using clinical-only features. The included clinical variables were sex, age, *NF1* disease, tumor location, extent of tumor resection, chemotherapy, and radiation. The forest plot of the trained Cox-PH model, and Kaplan-Meier curves of the stratified risk groups, as well as performance metrics on a discovery set of 160 patients are illustrated in the following figures and table.

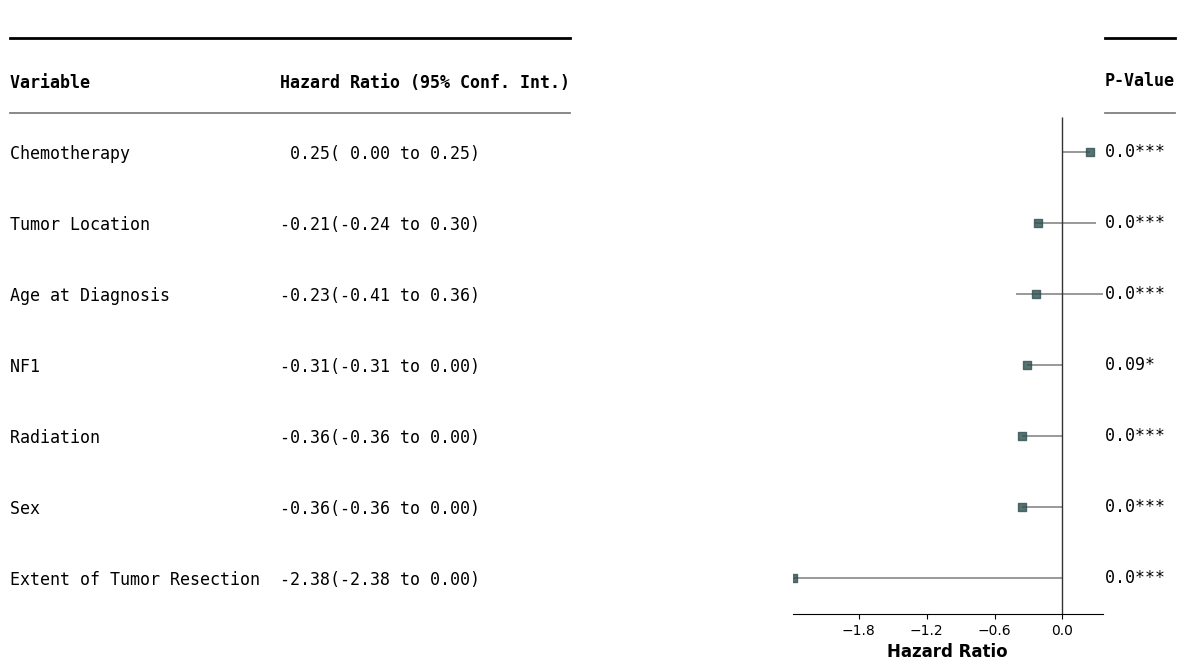

Supplemental Figure 7. Forest plot of the features in the clinical-only model.

| 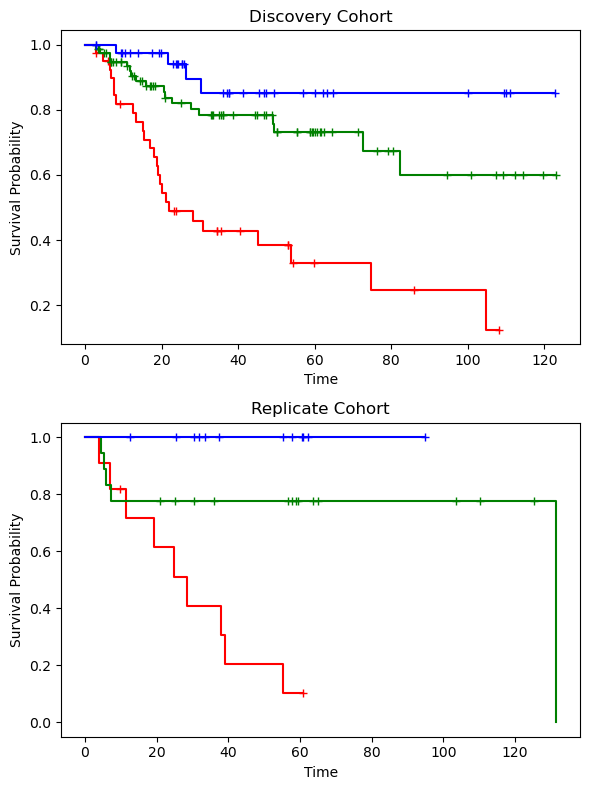 | 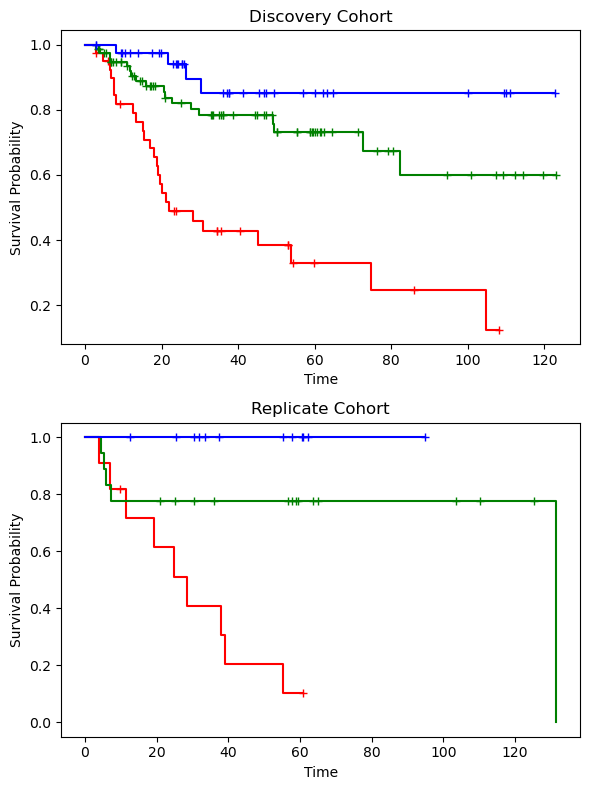 |
| --- | --- |

Supplemental Figure 8. Kaplan-Meier curves illustrating the risk categories for the model for clinical-only features and molecular subtypes.

Supplemental Table 3. Model performance metrics for the clinical-only model.

| **Cohort** | **HAR** | **UNO** | **IBS** |
| --- | --- | --- | --- |
| Discovery | 0.74 [95% CI: 0.67, 0.80] | 0.75 [95% CI: 0.67, 0.82] | 0.20 [95% CI: 0.16, 0.22] |
| Replicate | 0.76 | 0.79 | 0.16 |

##### 6.3.2 Model 2: Clinical Variables + Radiomics: Clinicoradiomic Model

We further investigated the performance of a Cox-PH model trained on a combination of clinical and radiomic features. As described in the Methods section, the clinical variables were not penalized, but regularization was applied to the radiomic feature set to reduce the dimensionality. The following plots and table depict the selected features for this model, Kaplan-Meier curves of the stratified risk groups and performance metrics. While the performance of the clinicoradiomic model on the unseen replication set was approximately similar to that of clinical-only model, the risk groups, especially the low-risk group from the medium and high-risk categories, are better differentiated as evidenced by the ANOVA p-value.

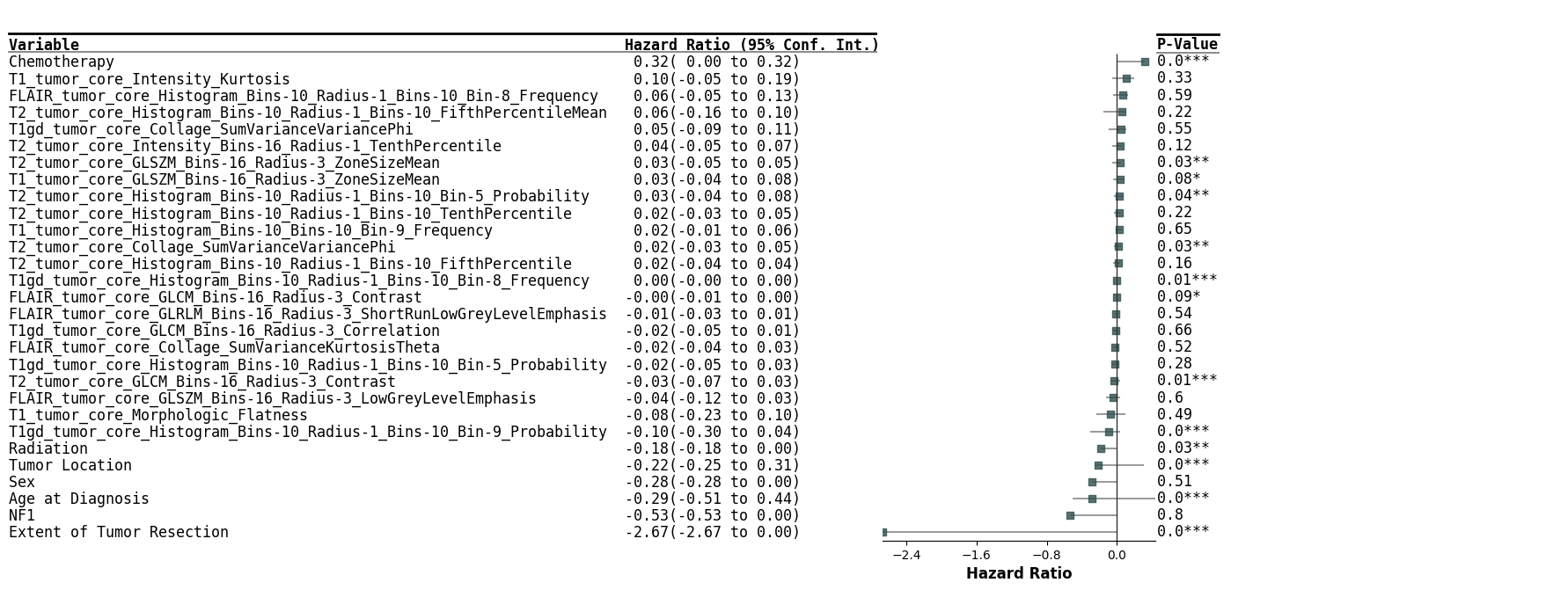

Supplemental Figure 9. Forest plot of the features in the clinicoradiomic model.

| 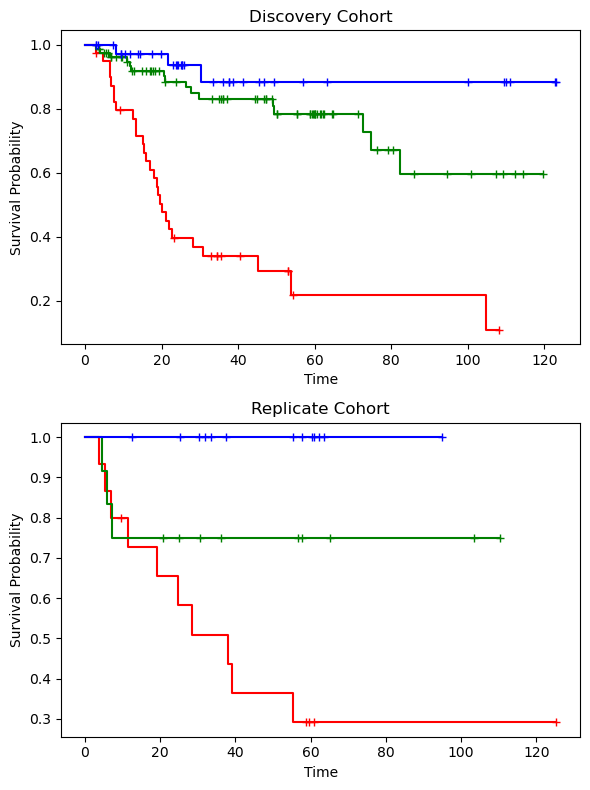 | 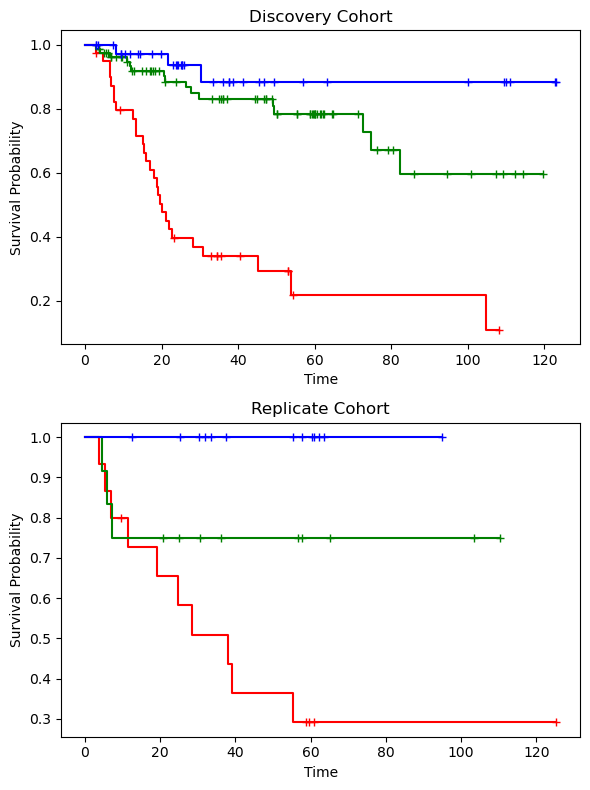 |
| --- | --- |

Supplemental Figure 10. Kaplan-Meier curves illustrating the risk categories for the model for clinicoradiomic features and molecular subtypes.

Supplemental Table 4. Model performance metrics for the clinicoradiomic model.

| **Cohort** | **HAR** | **UNO** | **IBS** |
| --- | --- | --- | --- |
| Discovery | 0.71 [95% CI: 0.63, 0.79] | 0.72 [95% CI: 0.64, 0.80] | 0.24 [95% CI: 0.19, 0.29] |
| Replicate | 0.77 | 0.80 | 0.16 |

***Comparative Analysis of Clinicoradiomic and Clinical-Only Models***

The forest plots for the clinical-only (Supplemental Figure 6) and clinicoradiomic (Supplemental Figure 8) models demonstrate that the extent of tumor resection is a critical predictive factor for tumor progression in pLGG, with patients undergoing gross/near-total resection exhibiting the most favorable prognosis. To evaluate the effectiveness of our clinicoradiomic model versus the clinical-only model in patients who did not receive gross/near-total resection, we conducted a focused subgroup analysis. This involved assessing the performance of the previously described clinical-only and clinicoradiomic models (Models 1 and 2, respectively) exclusively in patients who had not undergone gross/near-total tumor resection, intentionally excluding those who had. The results indicate that the clinicoradiomic model surpasses the clinical-only model in predicting progression-free survival for these patients.

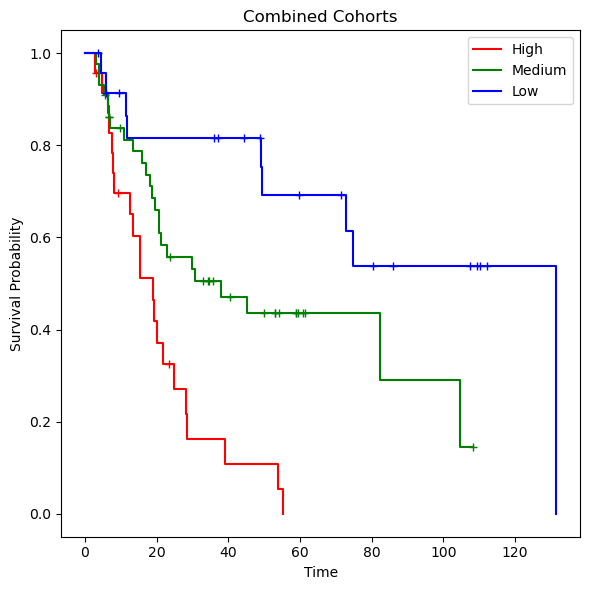

Supplemental Figure 11. Kaplan-Meier curves illustrating the risk categories for the model for clinicoradiomic features and molecular subtypes specifically on the subjects who did not undergo total/near-total resection.

Supplemental Table 5. Comparison of the performances of Model 1 (clinical-only) and Model 2 (clinicoradiomic) in a subgroup of patients who did not receive gross/near-total resection.

| **Model** | **HAR** | **UNO** | **IBS** |
| --- | --- | --- | --- |
| Model 1: Clinical-only | 0.55 | 0.57 | 0.24 |
| Model 2: Clinicoradiomic | 0.70 | 0.70 | 0.18 |

##### 6.3.3 Model 3: Clinical Variables + Radiomics + Molecular Subtypes

We further explored the predictive power of adding molecular subtypes, as a categorical variable, to the clinicoradiomic model. The performance of this model on the replication set is lower than the clinicoradiomic model, possibly due to smaller training cohort size.

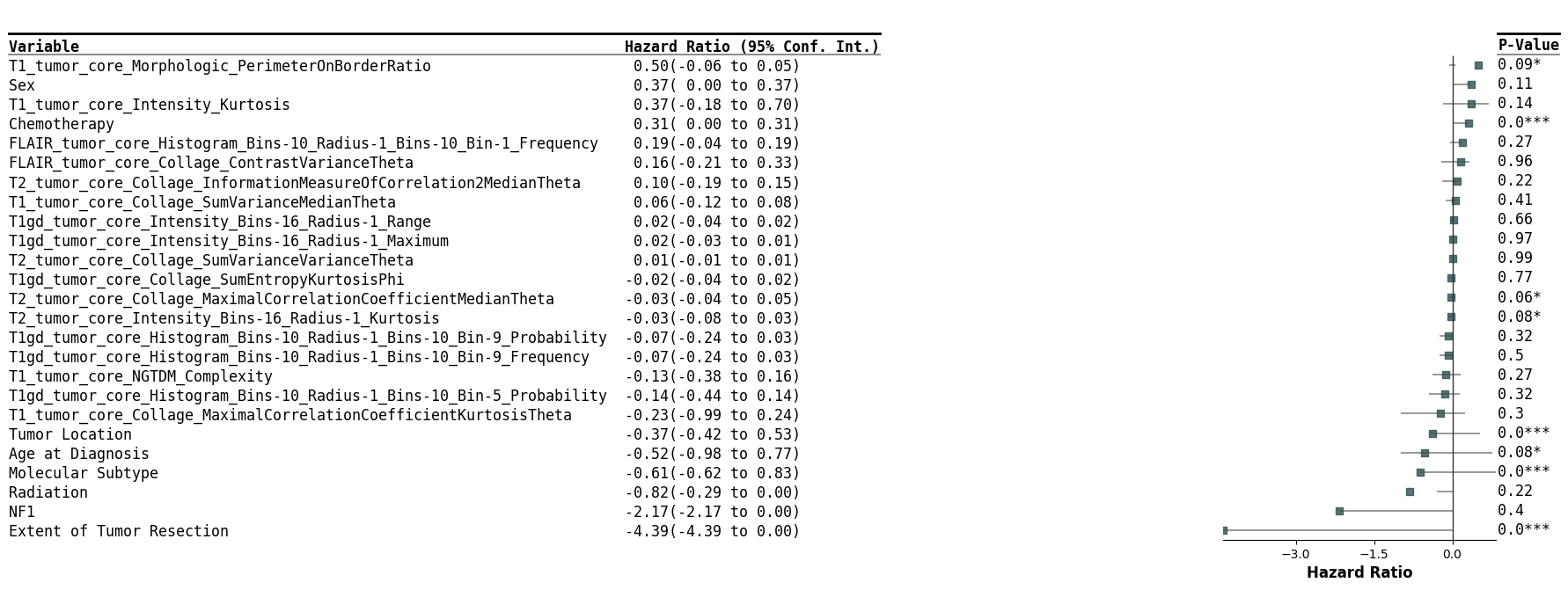

Supplemental Figure 12. Forest plot of the model comprised of clinicoradiomic features and molecular subtypes.

| 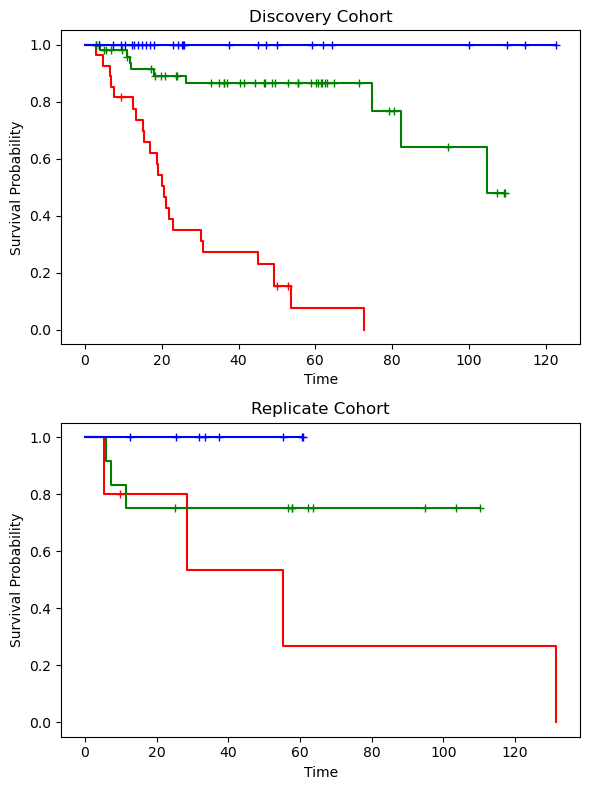 | 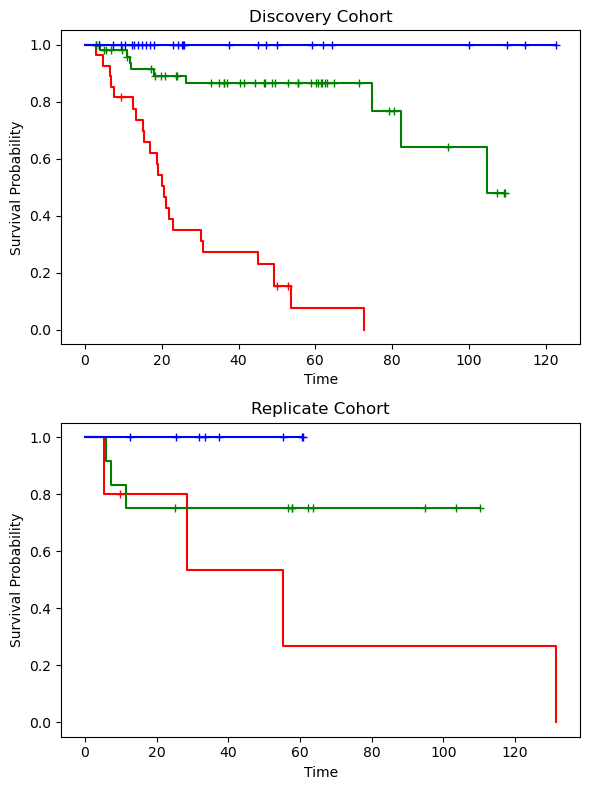 |
| --- | --- |

Supplemental Figure 13. Kaplan-Meier curves illustrating the risk categories for the model for clinicoradiomic features and molecular subtypes.

Supplemental Table 6. Model performance metrics for a combination of clinicoradiomic features and molecular subtypes.

| **Cohort** | **HAR** | **UNO** | **IBS** |
| --- | --- | --- | --- |
| Discovery | 0.74 [95% CI: 0.65, 0.83] | 0.74 [95% CI: 0.65, 0.84] | 0.21 [95% CI: 0.14, 0.27] |
| Replicate | 0.69 | 0.74 | 0.18 |
